# Brain-Computer Interface Robotics for Hand Rehabilitation After Stroke: A Systematic Review

**DOI:** 10.1101/2019.12.11.19014571

**Authors:** Paul Dominick E Baniqued, Emily C Stanyer, Muhammad Awais, Ali Alazmani, Andrew E Jackson, Mark A Mon-Williams, Faisal Mushtaq, Raymond J Holt

**Affiliations:** School of Mechanical Engineering, University of Leeds, LS2 9JT Leeds, United Kingdom; School of Psychology, University of Leeds, LS2 9JZ Leeds, United Kingdom

**Keywords:** brain-computer interface, EEG, robotics, rehabilitation, motor imagery, stroke

## Abstract

**Background:** Hand rehabilitation is core to helping stroke survivors regain activities of daily living. Recent studies have suggested that the use of electroencephalography-based brain-computer interfaces (BCI) can promote this process. Here, we report the first systematic examination of the literature on the use of BCI-robot systems for the rehabilitation of fine motor skills associated with hand movement and profile these systems from a technical and clinical perspective.

**Methods:** A search for January 2010-October 2019 articles using Ovid MEDLINE, Embase, PEDro, PsycINFO, IEEE Xplore and Cochrane Library databases was performed. The selection criteria included BCI-hand robotic systems for rehabilitation at different stages of development involving tests on healthy participants or people who have had a stroke. Data fields include those related to study design, participant characteristics, technical specifications of the system, and clinical outcome measures.

**Results:** 30 studies were identified as eligible for qualitative review and among these, 11 studies involved testing a BCI-hand robot on chronic and subacute stroke patients. Statistically significant improvements in motor assessment scores relative to controls were observed for three BCI-hand robot interventions. The degree of robot control for the majority of studies was limited to triggering the device to perform grasping or pinching movements using motor imagery. Most employed a combination of kinaesthetic and visual response via the robotic device and display screen, respectively, to match feedback to motor imagery.

**Conclusion:** 19 out of 30 studies on BCI-robotic systems for hand rehabilitation report systems at prototype or pre-clinical stages of development. We identified large heterogeneity in reporting and emphasise the need to develop a standard protocol for assessing technical and clinical outcomes so that the necessary evidence base on efficiency and efficacy can be developed.

## Background

There is growing interest in the use of robotics within the field of rehabilitation. This interest is driven by the increasing number of people requiring rehabilitation following problems such as stroke (with an ageing population), and the global phenomenon of insufficient numbers of therapists able to deliver rehabilitation exercises to patients [1,2]. Robotic systems allow a therapist to prescribe exercises that can then be guided by the robot rather than the therapist. An important principle within the use of such systems is that the robots assist the patient to actively undertake a prescribed movement rather than the patient’s limb being moved passively. This means that it is necessary for the system to sense when the patient is trying to generate the required movement (given that, by definition, the patient normally struggles with the action). One potential solution to this issue is to use force sensors that can detect when the patient is starting to generate the movement (at which point the robot’s motors can provide assistive forces). It is also possible to use measures of muscle activation (EMGs) to detect the intent to move [3]. In the last two decades there has been a concerted effort by groups of clinicians, neuroscientists and engineers to integrate robotic systems with brain signals correlated with a patient trying to actively generate a movement, or imagine a motor action, to enhance the efficacy and effectiveness of stroke rehabilitation-these systems fall under the definition of Brain Computer Interfaces, or BCIs [4].

BCIs allow brain state-dependent control of robotic devices to aid stroke patients during upper limb therapy. While BCIs in their general form have been in development for almost 50 years [5] and were theoretically made possible since the discovery of the scalp-recorded human electroencephalogram (EEG) in the 1920s [6], their application to rehabilitation is more recent [7–9]. Graimann et al. [10] defined a BCI as an artificial system that provides direct communication between the brain and a device based on the user’s intent; bypassing the normal efferent pathways of the body’s peripheral nervous system. A BCI recognises user intent by measuring brain activity and translating it into executable commands usually performed by a computer, hence the term “brain-computer interface”.

Most robotic devices used in upper limb rehabilitation exist in the form of exoskeletons or end-effectors. Robotic exoskeletons (i.e., powered orthoses, braces) are wearable devices where the actuators are biomechanically aligned with the wearer’s joints and linkages; allowing the additional torque to provide assistance, augmentation and even resistance during training [11]. In comparison, end-effector systems generate movement through applying forces to the most distal segment of the extremity via handles and attachments [11]. Rehabilitation robots are classified as Class II-B medical devices (i.e., a therapeutic device that administers the exchange of energy, mechanically, to a patient) and safety considerations are important during development [12,13]. Most commercial robots are focused on arms and legs, each offering a unique therapy methodology. There is also a category of device that target the hand and finger. While often less studied than the proximal areas of the upper limb, hand and finger rehabilitation are core component in regaining activities of daily living (ADL) [14]. Many ADLs require dexterous and fine motor movements (e.g. grasping and pinching) and there is evidence that even patients with minimal proximal shoulder and elbow control can regain some hand capacity long-term following stroke [15].

The strategy of BCI-robot systems (i.e. systems that integrate BCI and robots into one unified system) in rehabilitation is to recognise the patient’s intention to move or perform a task via a neural or physiological signal, and then use a robotic device to provide assistive forces in a manner that mimics the actions of a therapist during standard therapy sessions [16]. The resulting feedback is patient-driven and is designed to aid in closing the neural loop from intention to execution. This process is said to promote use-dependent neuroplasticity within intact brain regions and relies on the repeated experience of initiating and achieving a specified target [17,18]; making the active participation of the patient in performing the therapy exercises an integral part of the motor re-learning process [19,20].

The aforementioned scalp-recorded EEG signal is a commonly used instrument for data acquisition in BCI systems because it is non-invasive, easy to use and can detect relevant brain activity with high temporal resolution [21,22]. In principle, the recognition of motor imagery (MI), the imagination of movement without execution, via EEG can allow the control of a device independent of muscle activity [10]. It has been shown that MI-based BCI can discriminate motor intent by detecting event-related spectral perturbations (ERSP) [23,24] and/or event-related desynchronisation/synchronisation (ERD/ERS) patterns in the µ (9-11 Hz) and β (14-30 Hz) sensorimotor rhythm of EEG signals [24]. However, EEG also brings with it some challenges. These neural markers are often concealed by various artifacts and may be difficult to recognise through the raw EEG signal alone. Thus, signal processing (including feature extraction and classification) is a vital part of obtaining a good MI signal for robotic control. A general pipeline for EEG data processing involves several steps. First, the data undergo a series of pre-processing routines (e.g., filtering and artifact removal) before feature extraction and classification for use as a control signal for the robotic hand. There are variety of methods to remove artifact from EEG and these choices depend on the overall scope of the work [25]. For instance, Independent Component Analysis (ICA) and Canonical Correlation Analysis (CCA) can support real-time applications but are dependent on manual input. In contrast, regression and wavelet methods are automated but support offline applications. There also exist automated and real-time applications such as adaptive filtering or using blind source separation (BSS) based methods. Recently, the research community has been pushing real-time artifact rejection by reducing computational complexity e.g. Enhanced Automatic Wavelet-ICA (EAWICA) [26], hybrid ICA - Wavelet transform technique (ICA-W) [27] or by developing new approaches such as adaptive de-noising frameworks [28] and Artifact Subspace Reconstruction (ASR) [29]. Feature extraction involves recognising useful information (e.g., spectral power, time epochs, spatial filtering) for better discriminability among mental states. For example, the common spatial patterns (CSP) algorithm is a type of spatial filter that maximises the variance of band pass-filtered EEG from one class to discriminate it from another [30]. Finally, classification (which can range from linear and simple algorithms such as Linear Discriminant Analysis (LDA), Linear Support Vector Machine (L-SVM) up to more complex techniques in deep learning such as Convolutional Neural Networks (CNN) and Recurrent Neural Networks (RNN) [31,32] involves the translation of these signals of intent to an action that provides the user feedback and closes the loop of the motor intent-to-action circuit.

The potential of MI-based BCIs has gained considerable attraction because the neural activity involved in the control of the robotic device may be a key component in the rehabilitation itself. For example, MI of movement is thought to activate some of the neural networks involved in movement execution (ME) [33–36]. The resulting rationale is that encouraging the use of MI could increase the capacity of the motor cortex to control major muscle movements and decrease the necessity to use neural circuits damaged post-stroke. The scientific justification for this approach was first provided by Jeannerod [36] who suggested that the neural substrates of MI are part of a shared network that is also activated during the simulation of action by the observation of action (AO) [36]. These ‘mirror neuron’ systems are thought to be an important component of motor control and learning [36] - hence the belief that activating these systems could aid rehabilitation. The use of a MI-BCI to control a robot in comparison to traditional MI and physical practice provides a number of benefits to its user and the practitioner. These advantages include the fact that the former can provide a more streamlined approach such as sensing physiological states, automating visual and/or kinaesthetic feedback and enriching the task and increasing user motivation through gamification. There are also general concerns around the utility of motor imagery without physical movement (and the corresponding muscle development that comes from these) and it is possible that these issues could be overcome through a control strategy that progressively reduces the amount of support provided by the MI-BCI system and encourages active motor control [37,38].

A recent meta-analysis of the neural correlates of action (MI, AO and ME) quantified ‘conjunct’ and ‘contrast’ networks in the cortical and subcortical regions [33]. This analysis, which took advantage of open-source historical data from fMRI studies, reported consistent activation in the premotor, parietal and somatosensory areas for MI, AO and ME. Predicated on such data, researchers have reasoned that performing MI should cause activation of the neural substrates that are also involved in controlling movement and there have been a number of research projects that have used AO in combination with MI in neurorehabilitation [39–41] and motor learning studies [42,43] over the last decade.

One implication of using MI and AO to justify the use of BCI approaches is that great care must be taken with regard to the quality of the environment in which the rehabilitation takes place. While people can learn to modulate their brain rhythms without using motor imagery and there is variability across individuals in their ability to imagine motor actions, MI-driven BCI systems require (by design at least) for patient to imagine a movement. Likewise, AO requires the patients to clearly see the action. This suggests that the richness and vividness of the visual cues provided is an essential part of an effective BCI system. It is also reasonable to assume that feedback is important within these processes and thus the quality of feedback should be considered as essential. Afterall, MI and AO are just tools to modulate brain states and the effectiveness of these tools vary from one stroke patient to another [44]. Finally, motivation is known to play an important role in promoting active participation during therapy [20,45]. Thus, a good BCI system should incorporate an approach (such as gaming and positive reward) that increases motivation. Recent advances in technology make it far easier to create a rehabilitation environment that provides rich vivid cues, gives salient feedback and is motivating. For example, the rise of immersive technologies, including virtual reality (VR) and augmented reality (AR) platforms [46,45,47], allows for the creation of engaging visual experiences that have the potential to improve a patient’s self-efficacy [48] and thereby encourage the patient to maintain the rehabilitation regime. One specific example of this is visually amplifying the movement made by a patient when the movement is of limited extent so that the patient can see their efforts are producing results [49].

In this review we set out to examine the literature to achieve a better understanding of the current value and potential of BCI-based robotic therapy with three specific objectives:

1. Identify how BCI technologies are being utilised in controlling robotic devices for hand rehabilitation. Our focus was on the study design and the tasks that are employed in setting up a BCI-hand robot therapy protocol.
2. Document the readiness of BCI systems. Because BCI for rehabilitation is still an emerging field of research, we expected that most studies would be in their proof-of-concept or clinical testing stages of development. Our purpose was to determine the limits of this technology in terms of: (a) resolution of hand MI detection and (b) the degree of robotic control.
3. Evaluate the clinical significance of BCI-hand robot systems by looking at the outcome measures in motor recovery and determine if a standard protocol exists for these interventions.

It is important to note that there have been several recent reviews exploring BCI for stroke rehabilitation. For example, Monge-Pereira et al. [50] compiled EEG-based BCI studies for upper limb stroke rehabilitation. Their systematic review (involving 13 clinical studies on stroke and hemiplegic patients) reported on research methodological quality and improvements in the motor abilities of stroke patients. Cervera et al. [51] performed a meta-analysis on the clinical effectiveness of BCI-based stroke therapy among 9 randomised clinical trials (RCT). McConnell et al. [52] undertook a narrative review of 110 robotic devices with brain-machine interfaces for hand rehabilitation post-stroke. These reviews, in general, have reported that such systems provide improvements in both functional and clinical outcomes in pilot studies or trials involving small sample sizes. Thus, the literature indicates that EEG-based BCI are a promising general approach for rehabilitation post-stroke. The current work complements these previous reports by providing the first systematic examination on the use of BCI-robot systems for the rehabilitation of fine motor skills associated with hand movement and profiling these systems from a technical and clinical perspective.

## Methods

### Protocol Registration

Details of the protocol for this systematic review were registered on the International Prospective Register of Systematic Reviews (PROSPERO) and can be accessed at www.crd.york.ac.uk/PROSPERO (ID: CRD42018112107).

### Search Strategy and Eligibility

An in-depth search of articles from January 2010 to October 2019 was performed on Ovid MEDLINE, Embase, PEDro, PsycINFO, IEEE Xplore and Cochrane Library. Only full-text articles published in English were selected for this review. Table 1 shows the combination of keywords used in the literature searching.

**Table 1.** Keyword Combinations.

| Set 1 (OR) |  | Set 2 (OR) |  | Set 3 (OR) |
| --- | --- | --- | --- | --- |
| Brain-computer interface/BCI |  | Stroke (rehabilitation/ |  | Robotic (exoskeleton/ |
| Electroencephalography/EEG |  | therapy/treatment/recovery) |  | orthosis) |
| Brain-machine interface/BMI |  | Motor (rehabilitation, |  | Powered (exoskeleton/ |
| Neural control interface | AND | therapy/treatment/recovery) | AND | orthosis) |
| Mind-machine interface |  | Neurorehabilitation |  | Robot |
|  |  | Neurotherapy |  | Device |
|  |  | Hand (rehabilitation/therapy/<br>recovery/exercises/movement) |  |  |

The inclusion criteria for the articles were: (1) publications that reported the development of an EEG-based BCI; (2) studies targeted towards the rehabilitation of the hand after stroke; (3) studies that involved the use of BCI and a robotic device (e.g., exoskeleton, end-effector type, platform-types, etc.); (4) studies that performed a pilot test on healthy participants or a clinical trial with people who have had a stroke. The articles were also screened for the following exclusion criteria: (1) studies that targeted neurological diseases other than stroke; (2) studies that used other intention sensing mechanisms (electromyography/EMG, electrooculography/EOG, non-paretic hand, other body parts, etc.).

Two authors performed independent screenings of titles and abstracts based on the inclusion and exclusion criteria. The use of a third reviewer was planned a priori in cases where a lack of consensus existed around eligibility. However, consensus was achieved from the first two authors during this stage. Full-text articles were then obtained, and a second screening was performed until a final list of studies was agreed to be included for data extraction.

### Data Extraction

The general characteristics of the study and their corresponding results were extracted from the full-text articles by the reviewers following the Preferred Reporting Items for Systematic Reviews and Meta-Analysis (PRISMA) checklist. Data fields were extracted and categorised as follows:

- Participant characteristics: sample population, healthy or stroke patients, handedness, age, sex, acute or chronic stroke classification, and mean duration since stroke
- Study design: general description of study design, experimental and control groups
- Task design: description of the task instructed, and stimuli presentation (cue and feedback modalities, i.e.: visual, kinaesthetic, auditory, etc.)
- Technical specifications of the system: EEG system used (including number of channels), robot device used (e.g. hand exoskeleton, end-effector, etc.), actuation mode, and control strategy
- Main outcomes of the study: clinical outcomes (for studies involving stroke patients), classification accuracies (participant, group and study-levels), other significant findings

This data extraction strategy allowed us to further evaluate the technology and clinical use of the BCI-robot systems used in this study.

### Technology Evaluation

#### EEG Acquisition

The signal acquisition element of an EEG-based BCI is critical to its success in recognising task-related intent. To better understand current practice, we gathered the type of electrode used (i.e., standard saline-soaked, gel or dry electrodes), the number of channels and its corresponding placement in the EEG cap. To illustrate where signals are recorded from, we plotted the frequency with which electrodes were used across studies on a topographical map using the 10-20 international electrode placement system.

#### Signal Processing

We evaluated the signal processing strategies used by each study looking specifically at the feature extraction and classification techniques within the data pipeline. For the studies that reported classification accuracies (i.e., comparing the predicted class against the ground truth), we were able to compare their results among the current state-of-the-art classification accuracies published in literature.

#### Robot-Assisted Rehabilitation

As the receiving end of the BCI pipeline and the provider of kinaesthetic feedback to the user, the robot-assisted device for hand rehabilitation plays a key role in providing the intervention in this therapy regimen. The robot components were evaluated based on their actuation type, targeted body-part (i.e., single-finger, multi-finger, whole hand), and control strategy. We also reported on commercially available systems, which having passed a series of regulatory processes making them fit for commercial use, were classified as gold standard devices.

#### Technological Readiness

We assessed the development stages of the system as a whole by performing a Technological Readiness Assessment (TRA). Using this strategy, we were able to determine the maturity of the systems through a Technology Readiness Level (TRL) scale of 1-9 and quantify its implementation in a research or clinical setting [56]. Since a BCI-robot for rehabilitation can be categorised as a Class II-B medical device we have adapted a customised TRL scale to account for these requirements [56]. The customised TRL accounts for prototype development and pilot testing in human participants (TRL 3), safety testing (TRL 4-5), and small scale (TRL 6) to large scale (TRL 7-8) clinical trials. Performing a TRA on each device should allow us to map out where the technology is in terms of adoption and perceived usefulness. For example, if most of the studies have used devices that have TRL below the clinical trials stage (TRL 6-8), then we can have some confidence that said BCI-robot system is not yet widely accepted in the clinical community. In this way we can focus on questions that improve our understanding on the factors that impede its use as a viable therapy option for stroke survivors.

### Clinical Use

#### Clinical Outcomes Measures

For studies involving stroke patients, clinical outcomes were obtained based on muscle improvement measures such as Fugl-Meyer Motor Assessment Upper Extremity (FMA-UE) scores [53], Action Research Arm Test (ARAT) scores [54], United Kingdom Medical Research Council (UK-MRC) muscle grade [55], Grip Strength (GS) Test and Pinch Strength (PS) Test scores (i.e., kilogram force collected using an electronic hand dynamometer) among others.

#### Physiotherapy Evidence Database (PEDro) Scale for Methodological Quality

A methodological quality assessment was also performed for clinical studies based on the PEDro Scale [57]. This scale evaluates studies with a checklist of 11 items based on experts’ consensus criteria in physiotherapy practice. The complete details of the criteria can be found online [58]. A higher score in the PEDro scale (6 and above) implied better methodological quality but are not used as a measure of validity in terms of clinical outcomes. Pre-defined scores from this scale were already present in studies appearing in the PEDro search. However, studies without PEDro scores or are not present in the PEDro database at all had to be manually evaluated by the authors against the 11-item checklist (five of seven studies).

## Results

### Search Results

Figure 1 shows the study selection process and the number of articles obtained at each stage.

**Figure 1.**
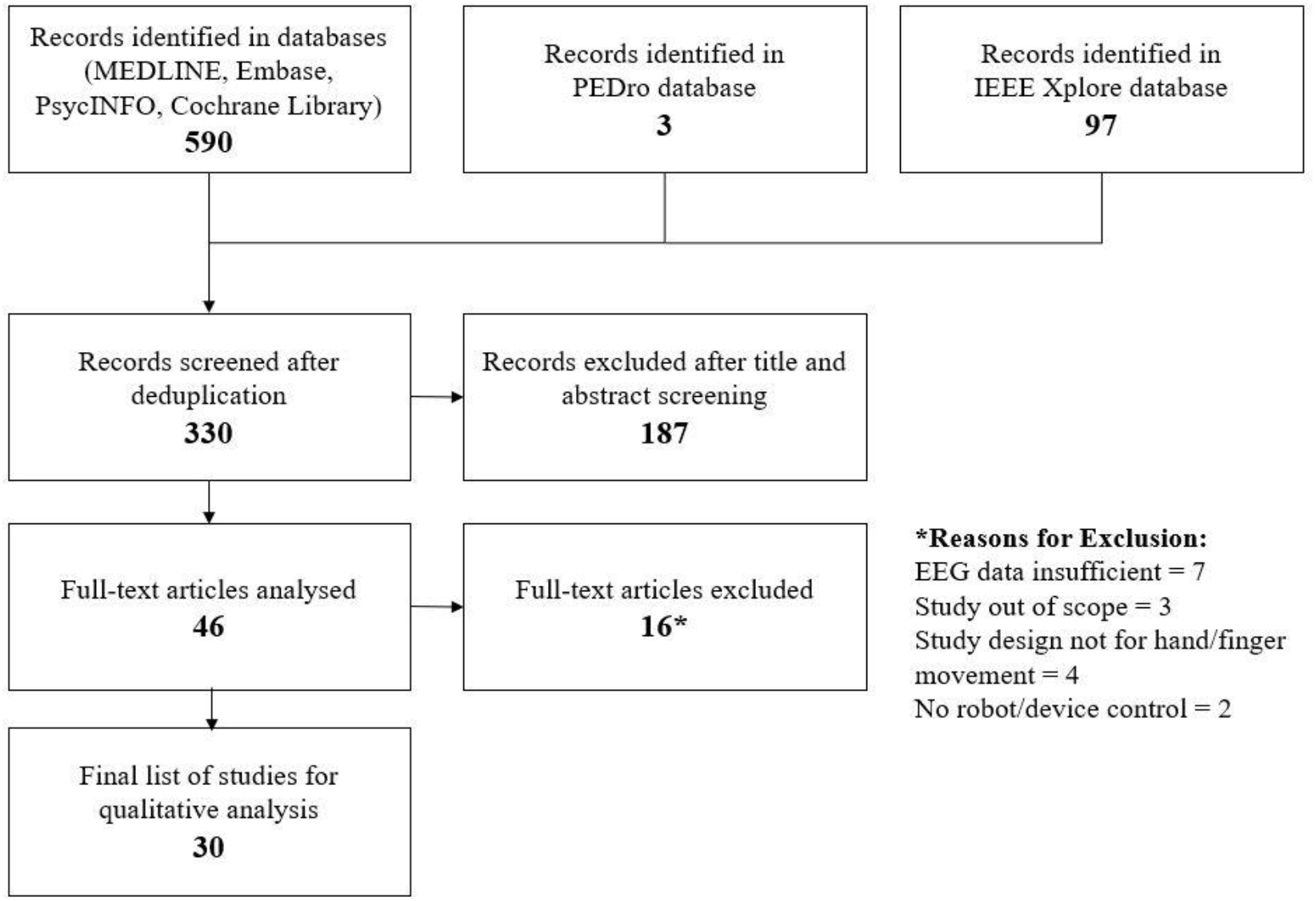
Study Selection Flowchart

A total of 590 studies were initially identified. After deduplication, 330 studies underwent title and abstract screening. Forty six studies passed this stage and among these, 16 were removed after full-text screening due to the following reasons: insufficient EEG and robotic data [59–65], the study was out of scope [66–68], the study design was not for hand/finger movement [69–72], no robot or mechatronic device was involved in the study [73,74]. A final sample of 30 studies were included in the qualitative review. Among the 30 studies, 11 [75– 85] were involved in testing the BCI-hand robot system on chronic and subacute stroke patients ([75,80] are RCTs) while the rest involved testing on healthy participants [86–104]. Table 2 shows a summary of the relevant data fields extracted from these studies.

**Table 2.** Summary of Studies.

| Authors | Participants | Study Design | Task Design | BCI-Hand Robot | Main Outcomes |
| --- | --- | --- | --- | --- | --- |
| **Studies involving stroke patients** |  |  |  |  |  |
| Ang et al., 2014 [75] | N=27 (7F:14M)<br>Moderate to severe impairment of UE function<br>Mean age: 54.2y<br>Mean stroke duration: 385.1 days | 3-armed RCT of motor function with MI-BCI-device as intervention<br>Control groups: device only (Haptic Knob), SAT | Photo manipulation: hand opening and closing, pronation and supination<br>Cue: visual (photo)<br>Feedback: visual (photo) and kinaesthetic | EEG: 27 channels to classify ERD/ERS and coupled with EMG to confirm MI<br>Device: Haptic Knob, 2-DOF for hand grasping and knob manipulation<br>Actuation: DC brushed motors with linear belt drive<br>Control: trigger | Clinical outcome measure: FMMA Distal, improvement in weeks 3, 6, 12, 24<br>BCI-device group = 2.5±2.4, 3.3±2.3, 3.2±2.7, 4.2±3.1<br>Device only group = 1.6±2.5, 2.9±3.0, 2.5±2.6, 2.5±3.0<br>SAT group = 0.4±1.1, 1.9±1.9, 1.0±1.3, 0.3±2.1 |
| Barsotti et al., 2015 [76] | N=3 (1F:2M)<br>Chronic stroke survivors with right arm hemiparesis<br>Mean age: 62±12y | Probing MI classification by BCI training, time-frequency analysis and robot trajectories<br>Uncontrolled | Reaching-grasping-releasing<br>Cue: visual<br>Feedback: kinaesthetic<br>Minimum time required to perform MI = 2s | EEG: 13 channels to classify ERD<br>Device: BRAVO 2-DOF hand orthosis attached to full UE exoskeleton<br>Actuation: DC motors with rigid links<br>Control: trigger | Mean classification accuracy during BCI training = 82.51±2.04%<br>Average delay from visual cue to robot initiation = 3.45±1.6s<br>Average delay due to patient's ability to start MI = 1.45s |
| Bundy et al., 2017 [77] | N=10<br>Chronic hemiparetic stroke with moderate to severe UE hemiparesis<br>Mean age: 58.6±10.3y | Motor function evaluation before and after intervention by MI-BCI from unaffected hemisphere | Opening of affected hand<br>Cue: visual<br>Feedback: visual and kinaesthetic | EEG: 8 channels to classify ERD<br>Device: 3-pinch grip, 1-DOF hand exoskeleton | Clinical outcome measure: ARAT Score, improvement from baseline to completion (12 weeks)<br>Mean ± SD = 6.20±3.81 |
| Bundy et al. (cont'd) |  | Uncontrolled |  | Control: continuous depending on spectral power | Note: 5.7 ARAT Score is the minimal clinically important difference in chronic stroke survivors |
| Carino-Escobar et al. 2019 [85] | N=9 (4F:5M)<br>Subacute ischaemic stroke<br>Mean age: 59.9±2.8y<br>Mean stroke duration: 158(±74)-185(±73) days | Determine longitudinal ERD/ERS patters and functional recovery with BCI-robot<br>Uncontrolled | Extension-flexion of hand fingers<br>Cue: visual (Graz MI)<br>Feedback: visual and kinaesthetic | EEG: 11 channels to classify ERD/ERS<br>Device: hand finger orthosis<br>Actuation: DC motor with screw system for linear displacement, flexible links<br>Control: trigger | FMA-UE: N=3 reported equal or higher than 3 score gains, N=3 no score gains,<br>Mean longitudinal ERD/ERS: beta bands have higher association with time since stroke onset than alpha, and strong association with UL motor recovery |
| Chowdhury et al., 2018-b [78] | N=20<br>10 healthy and 10 hemiplegic stroke patients<br>Mean age (healthy, stroke): 41±9.21y, 47.5±14.23y | Probe non-adaptive classifier (NAC) vs. Covariate Shift adaptive classifier (CSAC) of MI in EEG<br>Control group: healthy participants | Extension-flexion of hand fingers<br>Cue: visual<br>Feedback: visual and kinaesthetic | EEG: 12 channels with EMG to classify ERD/ERS<br>Device: EMOHEX 3-finger, 3-DOF each, exoskeleton (thumb, index, middle)<br>Actuation: servomotors with rigid links<br>Control: trigger | Mean classification accuracies during BCI training:<br>Healthy group: calibration = 78.50±9.01%, NAC = 75.25±5.46%, CSAC = 81.50±4.89%<br>Patient group: calibration = 79.63±13.11%, NAC = 70.25±3.43%, CSAC = 75.75±3.92% |
| Chowdhury et al., 2018-c [79] | N=4 (2F:2M)<br>Chronic hemiplegic stroke patients, right-handed, left hand impaired<br>Mean age: 44.75±15.69y<br>Mean stroke duration: 7 ±1.15mo | Motor function evaluation by using active physical practice followed by MI-BCI-controlled device intervention<br>Uncontrolled | Extension-flexion of hand fingers<br>Cue: visual<br>Feedback: visual and kinaesthetic | EEG: 12 channels with force sensors to classify ERD/ERS<br>Device: EMOHEX 3-finger, 3-DOF each, exoskeleton (thumb, index, middle)<br>Actuation: servomotors with rigid links<br>Control: trigger | Classification accuracies of 4 participants: P01 = 81.45±8.12%, P02 = 70.21±4.43%, P03 = 76.88±4.49%, P04 = 74.55±4.35%<br>Clinical outcome measures: GS and ARAT Scores, improvement from baseline to completion (6 weeks)<br>GS scores: group mean difference = +6.38 kg, p=0.06 |
| Chowdhury et al., 2018-c (cont'd) |  |  |  |  | ARAT scores: group mean difference = +5.66, p<0.05 |
| Frolov et al., 2017 [80] | N=74 (26F:48M)<br>BCI 55: Control 19<br>Subacute or chronic stroke with mild to hemiplegic hand paresis, right-handed | Multi-centre RCT of MI-BCI-controlled hand exoskeleton<br>Control group: SHAM | 3 Tasks: (1) motor relaxation, (2) imagery of left-hand opening, (3) imagery of right-hand opening<br>Cue: visual<br>Feedback: visual and kinaesthetic | EEG: 30 channels to classify the three mental tasks by Bayesian classifier based on covariance matrices<br>Device: hand exoskeleton by Neurobotics, Russia<br>Actuation: pneumatic motors with spring flexors<br>Control: trigger | Mean classification accuracy during BCI training = 40.6%<br>Clinical outcome measures:<br>FMMA Distal and ARAT Scores, improvement in 10 days of training<br>FMMA Distal = 2.0, p<0.01 (BCI) and 1.0, p=0.046 (control)<br>ARAT Grasp = 3.0, p<0.01 (BCI) and 1.0, p=0.0394 (control)<br>ARAT Grip = 1.0, p<0.01 (BCI) and 1.0, p=0.045 (control)<br>ARAT Pinch = 1.0, p<0.01 (BCI) and 0.0, p=0.675 (control) |
| Norman et al., 2018 [82] | N=8 (All male)<br>Chronic cortical and subcortical single haemorrhagic or ischaemic stroke (at least 6 months)<br>Mean age: 59.5±11.8y | Implementation of sensorimotor rhythm (SMR) control on robot-assistive movement<br>Uncontrolled | Extension of hand finger<br>Cue: visual<br>Feedback: visual and kinaesthetic | EEG: 16 channels mapping SMR changes<br>Device: FINGER robot<br>Actuation: Linear servo-tube actuator with rigid links<br>Control: Visual - continuous (colour change respective to SMR), Robot - trigger | Mean classification accuracies:<br>8 participants: 83.1%, 76.3%, 73.3%, 68.2%, 74.5%, 86.5%, 47.9%, 40.0%<br>Box and blocks test (BBT):<br>At screening: mean score = 14.3±10.0, mean change after therapy = 4.3±4.5 (range 0-12). Higher score changes in participants who demonstrated SMR control but not significant (p=0.199) |
| Ono et al., 2016-a [81] | N=21 (9F:12M)<br>Chronic stroke patients with hemiplegic hands<br>Mean age: 57.9±2.4y | Probe congruent vs. incongruent MI feedback strategies<br>Control groups: congruent (synchronous proprioceptive and visual feedback) and | Grasping of a tennis ball with a hand<br>Cue: visual (video of hand performing action)<br>Feedback: visual and kinaesthetic | EEG: 9 channels to classify ERD<br>Device: Power Assist Hand - Team ATOM, Atsugi, Japan | Mean classification accuracies:<br>Congruent feedback = 56.8±5.2%, chance level=36.4±4.5%<br>Incongruent feedback = 40.0±3.5%, chance level 35.4±4.5% |

| Ono et al., 2016-a<br>(cont'd) |  | incongruent<br>(proprioceptive<br>feedback given 1s after<br>visual) |  | Actuation: pneumatic<br>motors with rigid links<br>Control: trigger |  |
| --- | --- | --- | --- | --- | --- |
| Tsuchimoto et al.,<br>2019 [84] | N=18 (3F:14M)<br>Chronic haemorrhagic<br>or ischaemic stroke<br>(from 2mo onwards)<br>Mean age: 58±10y | Implementation of MI-controlled robotic<br>orthosis as<br>neurofeedback<br>Control: SHAM | Extension of hand finger<br>Cue: unspecified<br>Feedback: kinaesthetic<br>and electrical stimulation | EEG: 5 channels to<br>classify MI<br>Device: robotic finger<br>orthosis<br>Actuation: servo<br>motors with rigid links<br>Control: trigger | Significant time-intervention interaction<br>in the ipsilesional sensorimotor cortex.<br>Higher coactivation of sensory and motor<br>cortices for neurofeedback group in the<br>ipsilesional sensorimotor cortices as<br>compared to SHAM |
| Wang et al., 2018<br>[83] | N=24 (4F:20M)<br>Chronic stroke patients<br>with paralysed hands<br>Mean age: 54±9y | Implementation of<br>action observation and<br>motor imagery<br>(AO+MI) with<br>kinaesthetic feedback<br>Control: SHAM | Hand grasping<br>Cue: visual (video of<br>hand action / textual cues<br>in SHAM group)<br>Feedback: visual and<br>kinaesthetic | EEG: 16 channels to<br>classify ERD<br>Device: robot hand<br>Control: Trigger | AO+MI with kinaesthetic feedback<br>group showed significant improvements<br>in FMA-UE across longitudinal<br>evaluation [$\chi^2(2) = 7.659$, $p = 0.022$], no<br>significant difference in SHAM group<br>[$\chi^2(2) = 4.537$, $p = 0.103$] |
| Authors | Participants | Study Design | Task Design | BCI-Hand Robot | Main Outcomes |
| **Studies involving healthy participants** |  |  |  |  |  |
| Bauer et al., 2015<br>[97] | N=20 (11F:9M)<br>Right-handed<br>Mean age: 28.5±10.5y | Study on MI as<br>compared to motor<br>execution (ME) using<br>BCI-device | Opening of left hand<br>Cue: auditory<br>Feedback: kinaesthetic | EEG: 31 channels to<br>detect ERD, with EMG<br>to classify MI from<br>execution and account<br>for tonic contraction<br>Device: Amadeo,<br>Tyromotion, Austria<br>Control:<br>discontinuation of ERD<br>stops finger extension | Principal component analyses (between<br>MI and execution) generated coefficients<br>for the visual (VIS) and kinaesthetic<br>(KIS) imagery scale, BCI-robot<br>performance (BRI), tonic contraction<br>task (MOC) and visuomotor integration<br>task (VMI). VIS and KIS yielded high<br>coefficients on MI while MOC and VMI<br>yield high coefficients on ME. BRI show<br>high coefficient yields on both MI and<br>ME. |
| Cantillo-Negrete et al., 2015 [86] | N=1 | Design and implementation of a MI-controlled hand orthosis | Extension-flexion of right-hand finger<br>Cue: visual (modified Graz)<br>Feedback: kinaesthetic | EEG: 11 channels to detect MI<br>Device: 1-DOF hand finger orthosis<br>Actuation: DC motor with screw system for linear displacement, flexible links<br>Control: trigger | Classification accuracy = 78% |
| Chowdhury et al., 2015-a [87] | N=6<br>Age range: 20-30y | Study of cortico-muscular coupling in robotic finger exoskeleton control | Extension-flexion of hand fingers<br>Cue: visual<br>Feedback: kinaesthetic | EEG: 10 channels with EMG to classify MI<br>Device: 3-finger, 3-DOF each, exoskeleton (thumb, index, middle)<br>Actuation: servomotors with rigid links<br>Control: trigger | Mean classification accuracies: passive execution = 69.17%, hand execution = 71.25%, MI = 67.92% |
| Coffey et al., 2014 [92] | N=3 (All male)<br>Right-handed<br>Age range: 24-28y | Design and implementation of a MI-controlled hand orthosis | Hand digit and wrist contraction and extension<br>Cue: visual (Graz MI)<br>Feedback: kinaesthetic | EEG: 27 channels to classify MI<br>Device: hand glove controlled by Arduino<br>Actuation: pneumatic<br>Control: trigger | Glove inflation-deflation cycle = 22s<br>Classification accuracies of 3 participants: A = 92.5%, B = 90.0%, C = 80.0% |
| Diab et al., 2016 [103] | N=5 | Design and implementation of EEG-triggered wrist orthosis with accuracy improvement | Hand opening and closing<br>Cue: verbal instruction<br>Feedback: kinaesthetic | EEG: 14 channels to detect hand movement-related EEG<br>Device: actuated Talon wrist orthosis<br>Actuation: linear<br>Control: trigger | Mean classification accuracies: simulation studies = 95%, online BCI training = 86% |
| Fok et al., 2011 [102] | N=4 | Design and implementation of a MI-controlled hand orthosis | Hand opening and closing<br>Cue: unspecified<br>Feedback: visual (cursor movement) and kinaesthetic | EEG: 14 channels to detect MI-related ERD<br>Device: actuated Talon wrist orthosis<br>Actuation: linear actuator<br>Control: trigger | EEG signals from imagined hand movement was correlated with the contralesional hemisphere and utilised to trigger the actuation of orthosis<br>ERD was detected from 12 Hz bin power of EEG during move condition |
| Li et al., 2019 [88] | N=14 (4F:10M)<br>Mean age: 23.8±0.89y | Design and implementation of an attention-controlled hand exoskeleton with rigid-soft mechanism | Hand grasping<br>Cue: visual (video of hand action)<br>Feedback: kinaesthetic | EEG: 3 channels to map signals relative to attention<br>Device: hand exoskeleton<br>Actuation: linear actuator with rigid-soft mechanism<br>Control: Trigger | Mean classification accuracy:<br>95.54% actuation success rate against the attention threshold |
| Holmes et al., 2012 [93] | N=6 (All male, young adults) | Design and implementation of a MI-controlled hand orthosis | Hand opening and closing<br>Cue: textual<br>Feedback: kinaesthetic | EEG: 14 channels to detect hand movement-related EEG<br>Device: ExoFlex Hand Exoskeleton controlled by Arduino<br>Actuation: linear actuator connected to chained links that flex<br>Control: trigger | Classification accuracies of 6 participants: T001 = 95%, T002 = 98%, D001 = 91%, U001 = 93%, E001 = 87%, E002 = 86% |
| King et al., 2011 [104] | N=1 (Female) 24y | Contralateral control of hand orthosis using EEG-based BCI | Right hand idling and grasping<br>Cue: textual | EEG: 63 channels to control contralateral hand movement<br>Device: hand orthosis | Offline classification accuracy = 95.3±0.6%, $p < 3.0866 \times 10^{-25}$ |
| King et al. (cont'd) | | | Feedback: visual and kinaesthetic | Actuation: servomotors attached to Bowden cables as tendons<br>Control: trigger | Average lag from voluntary contractions to BCI-robot control = $2.24 \pm 0.19$s (after 5 sessions) |
| Naros et al., 2016 [98] | N=32 (16F:16M)<br>Mean age: $25.9 \pm 0.5$y | 2x2 factorial design with parameters: adaptive classifier threshold and non-adaptive classifier threshold, contingent feedback and non-contingent feedback | Opening of right hand<br>Cue: auditory<br>Feedback: kinaesthetic | EEG: 32 channels to detect ERD, with EMG to classify MI (FC3, C3, CP3 used)<br>Device: Amadeo, Tyromotion, Austria<br>Control: trigger | Significant enhancement in group 1 (adaptive classifier + contingent feedback), $p=0.0078$<br>Significant reduction in group 4 (non-adaptive classifier + non-contingent feedback), $p=0.0391$<br>Motor performance improvement over baseline from first and last tasks, significant results:<br>Group 1 (adaptive classifier + contingent feedback), $p=0.0313$<br>Group 4 = (non-adaptive classifier + non-contingent feedback), $p=0.0411$ |
| Ono et al., 2018-b [100] | N=28<br>Right-handed except 1 | Implementation of an action observation strategy with visual and proprioceptive, or auditory feedback to MI<br>Control group: SHAM | Grasping of a tennis ball with a hand<br>Cue: visual (video of hand performing action)<br>Feedback: visual, kinaesthetic and auditory | EEG: 9 channels to classify ERD<br>Device: Power Assist Hand - Team ATOM, Atsugi, Japan<br>Actuation: pneumatic motors with rigid links<br>Control: trigger | AO+MI + proprioceptive and visual feedback:<br>Mean MI-ERD powers of correct feedback vs SHAM provide significant interaction, $F_{1,17}=6.618$, $p=0.020$ (6 days)<br>Statistically significant increase in MI-ERD power in correct feedback group over baseline, $p=0.012$ (6 days) |
| Stan et al., 2015 [94] | N=9 | Trigger a hand orthosis using a P300 speller BCI | Spell E (enable), A (hand opening) and B (hand closing) in P300 speller BCI to perform hang grasping, moving and releasing objects<br>Cue: textual (spelling) | EEG: 8 channels focusing on visual cortex<br>Device: hand orthosis<br>Actuation: 2 servomotors and | Mean classification accuracies: 100% (on 6<sup>th</sup> letter flash during calibration) |
| Stan et al. (cont'd) |  |  | Feedback: visual (textual) and kinaesthetic | current feedback circuitry<br>Control: trigger |  |
| Ramos-Murguialday et al., 2012 [95] | N=23<br>Mean age (contingent positive, contingent negative, SHAM): 26.6±4y, 26.5±5y, 26.2±2y | Probing MI with proprioceptive feedback<br>Experimental groups: contingent positive, contingent negative feedback<br>Control group: SHAM | 5 tasks: MI without direct control, MI with direct control, passive, active, rest<br>Cue: auditory<br>Feedback: visual and kinaesthetic | EEG: 61 channels with EMG to classify ERD/ERS<br>Device: hand orthosis<br>Actuation: DC motor M-28 with a worm gearhead and Bowden cables for each finger<br>Control: trigger | Contingent positive feedback provided higher BCI performance during MI without feedback than contingent negative and SHAM; and higher during MI with or without feedback as compared to rest |
| Ramos-Murguialday and Birbaumer, 2015 [96] | N=9<br>Right-handed<br>Mean age: 26.6±4y | Detect oscillatory signatures of motor tasks during EEG | 5 tasks: MI without direct control, MI with direct control, passive, active, rest<br>Cue: auditory<br>Feedback: visual and kinaesthetic | EEG: 61 channels with EMG to classify ERD/ERS<br>Device: hand orthosis<br>Actuation: DC motor M-28 with a worm gearhead and Bowden cables for each finger<br>Control: trigger | Significant change in power in all frequency ranges during MI with direct control before trial initiation<br>Kinaesthetic feedback increased significant changes in alpha and beta power; therefore, increasing BCI performance |
| Randazzo et al., 2018 [90] | N=9 (2F:7M)<br>Mean age: 23±5y | Design and implementation of a hand orthosis with testing of kinaesthetic effects in EEG | 4 tasks: rest (REST), exoskeleton-induced hand motions (EXO), MI of right hand (MI), exoskeleton-induced hand motions plus MI (MIEXO)<br>Cue: visual<br>Feedback: kinaesthetic | EEG: 16 channels to detect MI<br>Device: mano hand exoskeleton<br>Actuation: linear servomotors attached to Bowden cables as tendons<br>Control: passive (exoskeleton not dependent on MI to | Mean classification accuracies among groups:<br>(vs REST) MI = 63.02±5.91%, EXO = 69.64±5.74%, MIEXO = 72.19±6.57%<br>MIEXO vs EXO = 69.91±9.86%<br>Chance level at 95% confidence = 58% (N=50 trials) |
| Randazzo et al.<br>(cont'd) |  |  |  | move during MIEXO task) |  |
| Tacchino et al., 2017<br>[91] | N=8 (7F:1M)<br>Right-handed<br>Mean age: 26.3±1.9y | 2x2 factorial design<br>with parameters: glove,<br>no glove, active<br>movement, passive<br>movement | Opening and closing of<br>hand, 4 tasks: (A) glove<br>with active movement,<br>(B) glove with passive<br>movement, (C) no glove<br>with active movement,<br>(D) no glove and no<br>movement<br>Cue: auditory<br>Feedback: kinaesthetic | EEG: 19 channels with<br>EMG to detect<br>ERD/ERS (C3, F3, Cz<br>used)<br>Device: Gloreha hand<br>rehabilitation glove<br>Actuation: electric<br>actuators with Bowden<br>cables on each finger<br>Control: passive (glove<br>not dependent on brain-<br>state during tasks) | Statistically significant ERD changes in<br>beta and mu bands were observed to<br>initiate earlier in tasks A and C (involves<br>active movement)<br>Stronger and longer ERD was observed<br>in tasks A and B (involves robotic<br>assistance) suggesting reinforced afferent<br>kinaesthetic feedback |
| Vukelic and<br>Gharabaghi, 2015<br>[99] | N=11 (4F:7M)<br>Right-handed<br>Mean age: 25.83±3.1y | Assessment<br>sensorimotor activity<br>during MI with either<br>visual or kinaesthetic<br>feedback | Right hand opening<br>Cue: visual (coloured<br>cursor ball)<br>Feedback: visual and<br>kinaesthetic (separated<br>by experimental groups) | EEG: 128 channels to<br>detect ERD/ERS<br>during MI (F3, CP3,<br>C3 used)<br>Device: Amadeo,<br>Tyromotion, Austria<br>Control: trigger | MI + kinaesthetic feedback group<br>resulted in higher beta ERS (p=0.02)<br>during rest and higher beta ERD<br>(p=0.04) during MI<br>Kinaesthetic feedback provides higher<br>stability and sustained beta ERD activity<br>than visual feedback |
| Witkowski et al.,<br>2014 [101] | N=12 (4F:8M)<br>Right-handed<br>Mean age: 28.1±3.63y | Assessment<br>performance and safety<br>of EEG-EOG hybrid<br>BCI | Right hand grasping<br>Cue: visual (coloured<br>squares and arrows)<br>Feedback: kinaesthetic | EEG: 5 channels with<br>EOG and EMG to<br>detect ERD during MI<br>Device: HX hand<br>exoskeleton<br>Actuation: DC motors<br>with Bowden cables for<br>thumb and index<br>fingers<br>Control: trigger | Mean classification accuracies:<br>EEG only = 63.59±10.81%<br>EEG/EOG hybrid = 60.77±9.42%<br>Mean safety criterion violations during<br>rest:<br>EEG only = 45.91±26.8%<br>EEG/EOG hybrid = 10.14±0.3% |
| Zhang et al., 2019<br>[89] | N=6 (2F:4M)<br>Right-handed<br>Age range: 23-26y | Implementation of a multimodal system using EEG, EMG and EOG to control a soft-robotic hand | Graz visualisation and auditory instructions, eye movements and physical practice (hand gestures)<br>Cue: visual (Graz MI), auditory<br>Feedback: visual and kinaesthetic | EEG with EMG and EOG: 40 channels to analyse ERD/ERS patterns<br>Device: Soft pneumatic finger<br>Actuation: pneumatic actuator with soft structures<br>Control: trigger | Mean classification accuracies:<br>EOG = 94.23%<br>EEG = 31.46%<br>EMG = 36.38%<br>Multimodal = 93.83±0.02% |

UE = Upper Extremity, MI = Motor Imagery, BCI = Brain-Computer Interface, RCT = Randomised Clinical Trial, SAT = Standard Arm Therapy, EMG = Electromyography, EOG = Electrooculography, ERD/ERS = Event-Related Desynchronisation/Synchronisation, FMMA = Fugl-Meyer Motor Assessment, ARAT = Action Research Arm Test, GS = Grip Strength, DOF = Degrees-of-Freedom

#### Studies with Healthy Participants (Prototype Group)

The studies which involved pilot testing on healthy human participants had a combined total of 207 individuals (sample size ranging from 1-32) who had no history of stroke or other neurological diseases. Right-handed individuals made up 44.24% of the combined population while the other 55.76% were unreported. These studies aimed to report the successful implementation of a BCI-robot system for hand rehabilitation and were more heterogeneous in terms of study and task designs than those studies that involved clinical testing. The most common approach was to design and implement a hand orthosis controlled by MI which accounted for 9 out of the 19 studies and were measured based on classification accuracy during the calibration/training period and online testing. Li et al. [88] and Stan et al. [94] also aimed to trigger a hand orthosis but instead of MI, the triggers used by Li et al. is based on an attention threshold while Stan et al. used a vision-based P300 speller BCI. Bauer et al. [97] compared MI against ME using a BCI-device while Ono et al. [100] studied the implementation of an action observation strategy with a combined visual and kinaesthetic feedback or auditory feedback. Five more studies [91,95,96,98,99] focused on varying the feedback while two more [89,101] assessed the performance and safety of a hybrid BCI with EMG, EOG or both.

#### Studies with Stroke Patients (Clinical Group)

A total of 208 stroke patients (with sample size varying 3-74) were involved in the 11 clinical studies. One study [75] reported a 3-armed RCT with control groups as device-only and SAT while another study [80] was a multi-centre RCT with sham as the control group. Five studies were uncontrolled – where the aims were either to study classification accuracies during sessions [76], to monitor clinical outcomes improvement from Day 0 until the end of the programme [77,85] or both [79,82]. Two studies [83,84] compared effects of the intervention against SHAM feedback. Another study [78] compared the classification accuracies of healthy and hemiplegic stroke patients against two BCI classifiers while the remaining study [81] compared classification accuracies from stroke patients who receive congruent or incongruent visual and kinaesthetic feedback.

### Technology Evaluation

#### EEG Acquisition

The EEG acquisition systems involved in the studies ranged from low-cost devices having few electrode channels (2-15 gel or saline-soaked silver/silver chloride [Ag/AgCl] electrodes) to standard EEG caps that had higher spatial resolution (16-256 gel or saline-soaked Ag/AgCl electrodes). The placement of EEG channels was accounted for by studies involving MI (N=21). This allowed us to determine the usage frequency among electrodes and is presented in Figure 2 as a heat map generated in R Studio (using the packages: “akima”, “ggplot2” and “reshape2”) against the 10-20 international electrode placement system.

**Figure 2.**
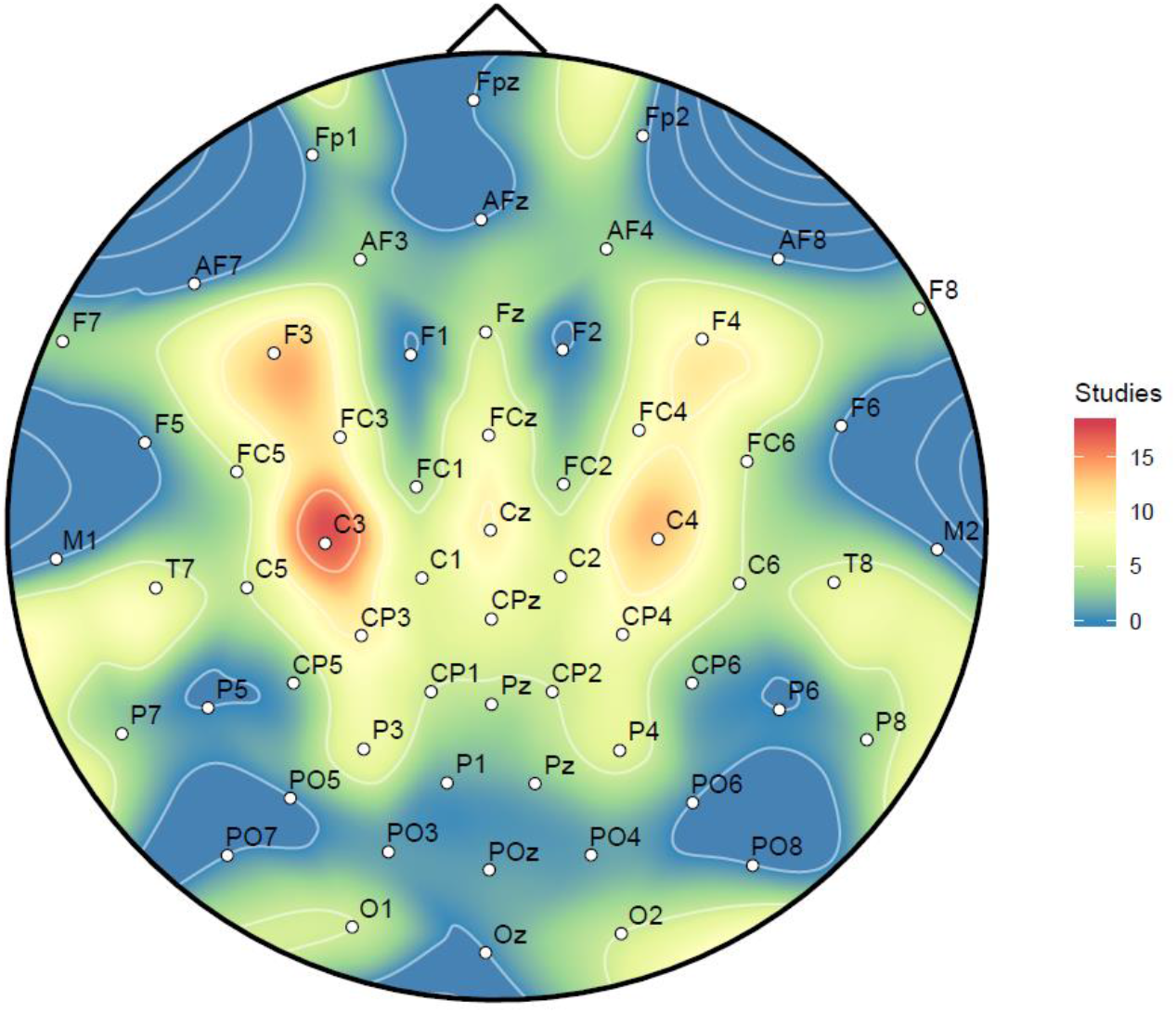
EEG Channel Usage across Motor Imagery Studies (N=21)

It can be seen that the EEG channels used for MI studies are concentrated towards electrodes along the central sulcus (C) region and the frontal lobe (F) region of the placement system where the motor cortex strip lies. Among these, C3 (N=17) and F3 (N=14) were mostly used, presumably because a majority of the participants were right-handed. The next most frequent were C4 (N=13) and the electrodes F4, Cz and CP3 (N=10).

#### Signal Processing: Feature Extraction and Classification

In the EEG-based BCI studies examined, it was found that the feature extraction and classification techniques were variable between systems. Table 3 provides a summary of pre-processing, feature extraction and classification techniques across the studies. There was a wide variation in the implemented signal processing strategies, but a unifying theme across studies was the attempt to: (i) discriminate mental states recorded in EEG across different manual tasks; (ii) classify the different states to produce a viable signal.

**Table 3.** BCI Feature Extraction and Classification.

| Study | Pre-Processing | Feature Extraction | Classification | Hand Task |
| --- | --- | --- | --- | --- |
| Ang et al. [75] | Band-pass<br>(0.05-40 Hz) | Filter Bank Common<br>Spatial Pattern<br>(FBCSP) algorithm<br>[105] | Calibration model<br>(unspecified) | MI vs rest |
| Barsotti et al. [76] | Band-pass (8-24<br>Hz) | ERD ($\beta$ and μ-<br>decrease), CSP | SVM with linear kernel | MI vs rest |
| Bauer et al. [97] | Band-pass (6-16<br>Hz using zero-<br>phase lag FIR | ERD (β-decrease) | Linear autoregressive model<br>based on Burg Algorithm | MI vs rest |
| Bundy et al. [77] | Unspecified | ERD ($\beta$ and μ-<br>decrease) | Linear autoregressive model | MI (affected,<br>unaffected) vs<br>rest |
| Chowdhury et al.<br>[78] | Band-pass (0.1<br>Hz-100 Hz),<br>Notch (50 Hz) | CSP Covariance-<br>based, ERD/ERS ($\beta$<br>and μ-change) | SVM with linear kernel,<br>Covariate Shift Detection<br>(CSD)-based Adaptive<br>Classifier | left vs right MI |
| Coffey et al. [92] | Band-pass (0.5<br>Hz-30 Hz),<br>Notch (50 Hz) | CSP Covariance-<br>based | Linear Discriminant<br>Analysis (LDA) classifier | MI vs rest |
| Diab et al. [103] | Unspecified | Time epochs<br>(unspecified) | Artificial Neural Network<br>(ANN)-based Feed Forward<br>Back Propagation | Non-MI open vs<br>closed |
| Frolov al. [80] | Band-pass (5-30<br>Hz), FIR (order<br>101), IIR notch<br>Chebyshev type<br>I filter (50 Hz) | Time epochs<br>(10 s) | Bayesian-based EEG<br>covariance classifier [106] | MI (affected,<br>unaffected) vs<br>rest |
| Ono et al. [81] | Band-pass (0.5-30 Hz), notch<br>(50 or 60 Hz) | Time epochs (700<br>ms), ERD (μ-<br>decrease) | Linear Discriminant<br>Analysis (LDA) classifier | MI vs rest |
| Ramos-Murguialday<br>et al. [95] | Unspecified | Time epochs (5 s),<br>Spatial filter,<br>ERD/ERS ($\beta$ and μ-<br>change) | Linear autoregressive model | MI vs rest |
| Vukelic and<br>Gharabaghi [99] | High-pass<br>(unspecified) | ERD (β-decrease) | Linear autoregressive model<br>based on Burg Algorithm | MI vs rest |
| Witkowski et al.<br>[101] | Band-pass (0.4-70 Hz),<br>Laplacian filter | ERD/ERS ($\beta$ and μ-<br>change) | Linear autoregressive model<br>based on Yule-Walker<br>algorithm | MI vs rest |

SVM = Support Vector Machines, FIR = Finite Impulse Response, IIR = Infinite Impulse Response

While classification accuracy is contingent on the number of mental state classes the system is trying to discriminate, classification accuracies do provide a comparable metric among BCI systems. In our review, we found high variation in the reported mean classification accuracies among the BCI systems in this study (i.e., 2-class left-hand and right-hand classification)-ranging from 40% (below chance-level) [80–82] up to 95% [88,93,103,104]. For reference, two recent reviews on the state-of-the-art in classification accuracies for motor imagery BCI find ranges between 63-97% [107] and 68-90% [108].

#### Robot-Assisted Rehabilitation

Robotic hand rehabilitation systems provide kinaesthetic feedback to the user during BCI trials. Most of these devices are powered by either DC motors, servomotors or pneumatic actuators that transmit energy via rigid links or Bowden cables in a tendon-like fashion. The studies in this review included single-finger [84–86], multi-finger [82] (including EMOHEX [78,79,87]), full hand gloves [88,89] (including mano: Hand Exoskeleton [90] and Gloreha [91]) and full arm exoskeletons with isolated finger actuation (BRAVO-Hand [76]). Nine of the studies [77,87,88,90,92–96] presented their novel design of a hand rehabilitation device within the article while some reported on devices reported elsewhere (i.e., in a previous study of the group or a research collaborator). Two commercially-available devices were also used: AMADEO (Tyromotion, Austria) is an end-effector device used in 3 studies [97–99], and Gloreha (Idrogenet, Italy) is a full robotic hand glove used by Tacchino et al. [91]. AMADEO and Gloreha are both rehabilitation devices that have passed regulatory standards in their respective regions. AMADEO remains the gold standard for hand rehabilitation devices as it has passed safety and risk assessments and provided favourable rehabilitation outcomes. The International Classification of Functioning, Disability and Health (ICF) provides three specific domains that can be used to assess an intervention of this kind: improving impairments, supporting performance of activities and promoting participation [109,110]. In this case, a gold standard device not only prioritises user safety (established early in the development process) but also delivers favourable outcomes in scales against these domains. Figure 3 shows the main types of robotic hand rehabilitation devices.

**Figure 3.**
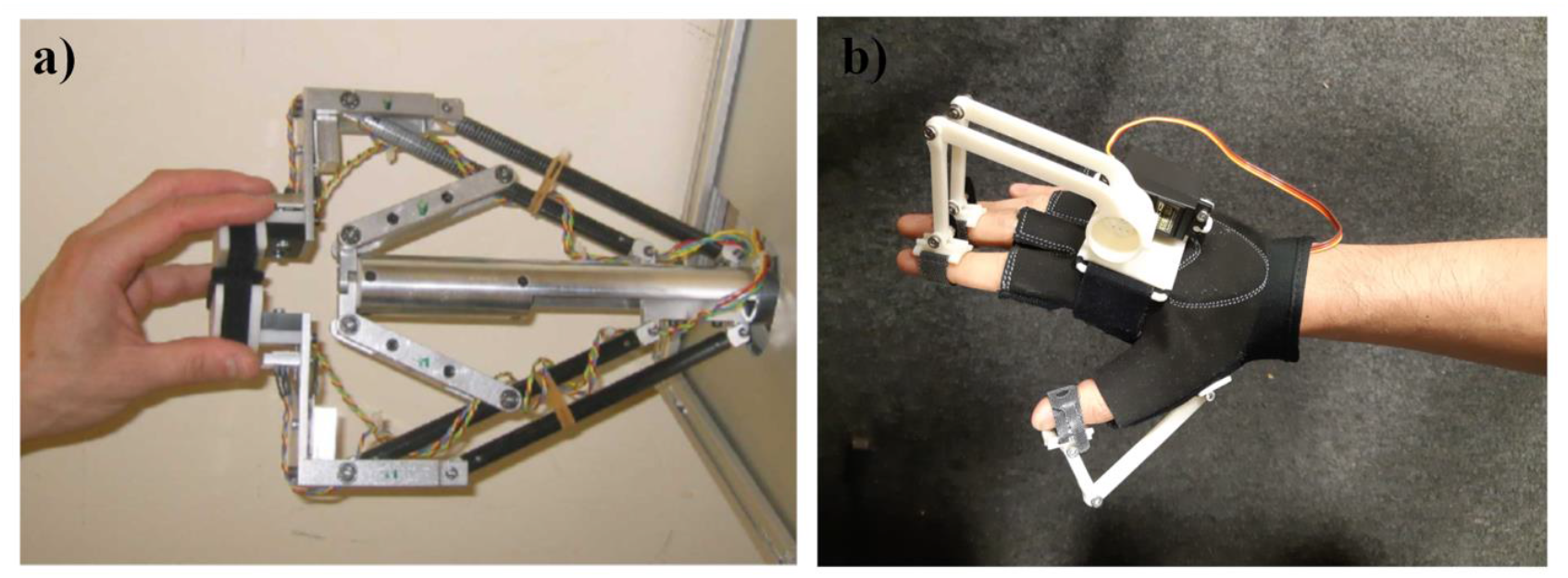
Robotic hand rehabilitation devices: a) An end-effector device (Haptic Knob) used in one of the extracted studies [75,111], b) a wearable hand exoskeleton/orthosis

#### Technology Readiness Assessment

A Technology Readiness Assessment (TRA) [56] was performed for each study and the Technology Readiness Levels (TRL) are presented in Table 4. While some of the system components (especially among robotic devices) were commercially available (having TRL 9+), we performed a TRA on the whole system (the interaction between BCI and robotics) to provide an evaluation of its maturity and state-of-the-art development with regard to rehabilitation medicine. We further assessed the TRL of each system at the time of the publication and its subsequent development.

**Table 4.**
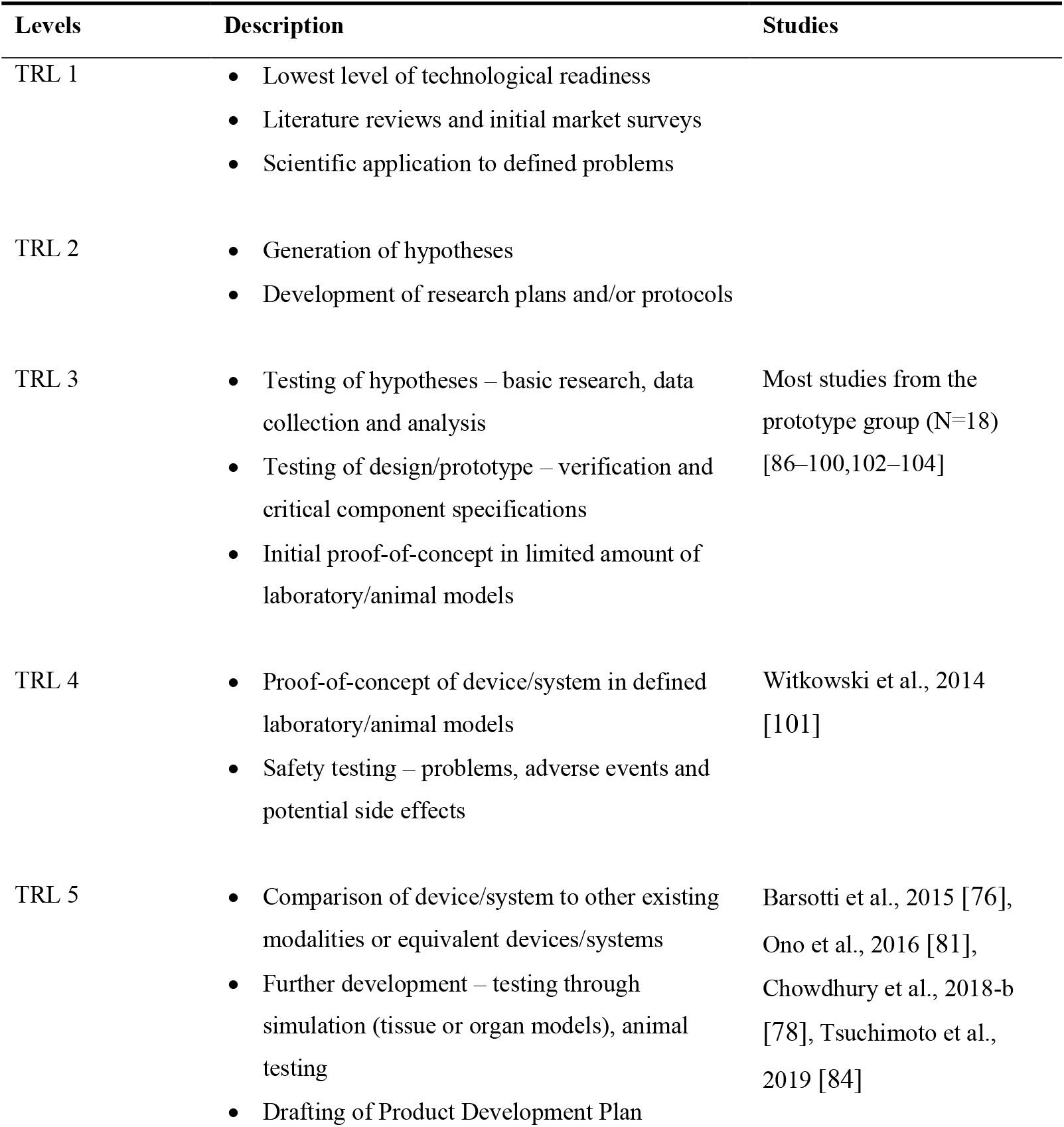
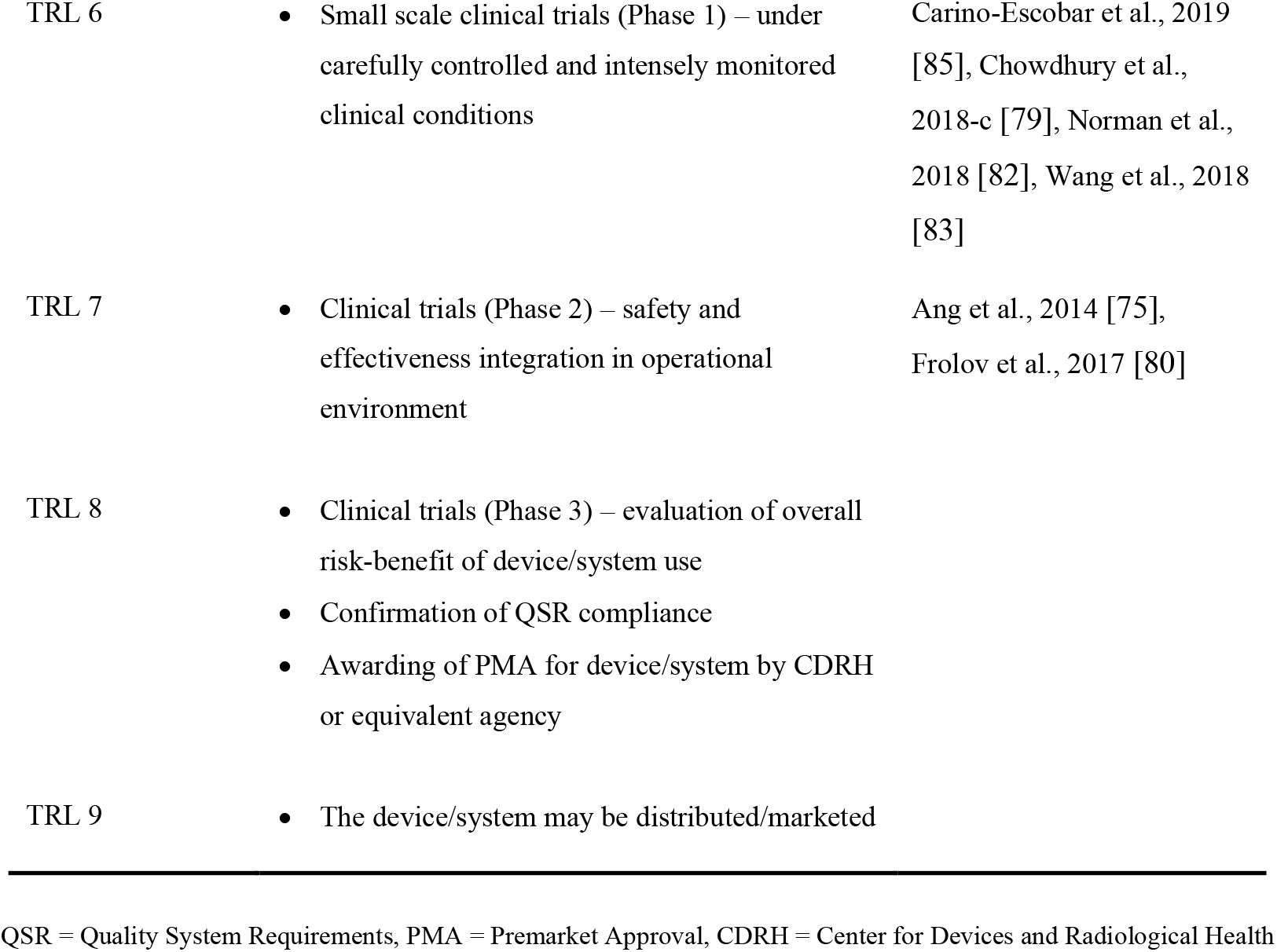
Technology Readiness Assessment of the BCI-Hand Robot Systems.

| Levels | Description | Studies |
| --- | --- | --- |
| TRL 1 | • Lowest level of technological readiness<br>• Literature reviews and initial market surveys<br>• Scientific application to defined problems |  |
| TRL 2 | • Generation of hypotheses<br>• Development of research plans and/or protocols |  |
| TRL 3 | • Testing of hypotheses – basic research, data collection and analysis<br>• Testing of design/prototype – verification and critical component specifications<br>• Initial proof-of-concept in limited amount of laboratory/animal models | Most studies from the prototype group (N=18) [86–100,102–104] |
| TRL 4 | • Proof-of-concept of device/system in defined laboratory/animal models<br>• Safety testing – problems, adverse events and potential side effects | Witkowski et al., 2014 [101] |
| TRL 5 | • Comparison of device/system to other existing modalities or equivalent devices/systems<br>• Further development – testing through simulation (tissue or organ models), animal testing<br>• Drafting of Product Development Plan | Barsotti et al., 2015 [76], Ono et al., 2016 [81], Chowdhury et al., 2018-b [78], Tsuchimoto et al., 2019 [84] |
| TRL 6 | • Small scale clinical trials (Phase 1) – under carefully controlled and intensely monitored clinical conditions | Carino-Escobar et al., 2019 [85], Chowdhury et al., 2018-c [79], Norman et al., 2018 [82], Wang et al., 2018 [83] |
| TRL 7 | • Clinical trials (Phase 2) – safety and effectiveness integration in operational environment | Ang et al., 2014 [75], Frolov et al., 2017 [80] |
| TRL 8 | • Clinical trials (Phase 3) – evaluation of overall risk-benefit of device/system use<br>• Confirmation of QSR compliance<br>• Awarding of PMA for device/system by CDRH or equivalent agency |  |
| TRL 9 | • The device/system may be distributed/marketed |  |

QSR = Quality System Requirements, PMA = Premarket Approval, CDRH = Center for Devices and Radiological Health

### Clinical Use

#### Clinical Outcomes Measures

Most of the studies adopted FMA-UE, ARAT and GS measurements to assess clinical outcomes. Six studies [75,77,79,80,83,85] reported patient improvement in these measures when subjected to BCI-hand robot interventions; in contrast with their respective controls or as recorded through time in the programme. For Ang et al. [75], FMA-UE Distal scores were reported in weeks 3, 6, 12 and 24 and the BCI-device group (N=6) yielded the highest improvement in scores across all time points as compared to the device only (N=8) and SAT (N=7) groups. Bundy et al. [77] reported an average of 6.20±3.81 improvement in the ARAT scores of its participants (N=10) in the span of 12 weeks while Chowdhury et al. [79] reported a group mean difference of +6.38 kg (p=0.06) and +5.66 (p<0.05) in GS and ARAT scores, respectively (N=4). Frolov et al.’s [80] multi-centre RCT reported a higher improvement in the FMA-UE Distal, ARAT Grasp and ARAT Pinch scores of the BCI-device group (N=55) when compared to the control/SHAM group (N=19), but not in the ARAT Grip scores where the values are both equal to 1.0 with p<0.01 for the BCI-device group and p=0.045 for the control.

#### Physiotherapy Evidence Database (PEDro) Scale for Methodological Quality

For the studies that had a clinical testing component, a methodological quality assessment by the PEDro Scale was performed. Two studies which appeared on the PEDro search [75,80] had predetermined scores in the scale and were extracted for this part while the rest were manually evaluated by the authors. Table 5 shows the results of the methodological quality assessment against the scale. Note that in the PEDro Scale, the presence of an eligibility criteria is not included in the final score.

**Table 5.** Methodological Quality of Clinical Studies based on PEDro Scale.

| Criteria |  | Ang et al. | Barsotti et al. | Bundy et al. | Carino-Escobar et al. | Chowdhury-b | Chowdhury-c | Frolov et al. | Norman et al. | Ono et al. | Tsuchimoto et al. | Wang et al. |
| --- | --- | --- | --- | --- | --- | --- | --- | --- | --- | --- | --- | --- |
| 1 | Eligibility criteria* | 1 | 1 | 1 | 1 | 1 | 1 | 1 | 1 | 0 | 1 | 1 |
| 2 | Random allocation | 1 | 0 | 0 | 0 | 0 | 0 | 1 | 0 | 0 | 1 | 1 |
| 3 | Concealed allocation | 0 | 0 | 0 | 0 | 0 | 0 | 0 | 0 | 0 | 1 | 1 |
| 4 | Baseline comparability | 1 | 0 | 1 | 0 | 1 | 1 | 1 | 0 | 0 | 0 | 0 |
| 5 | Blind subjects | 0 | 0 | 0 | 0 | 0 | 0 | 0 | 0 | 0 | 1 | 1 |
| 6 | Blind therapists | 0 | 0 | 0 | 0 | 0 | 0 | 0 | 0 | 0 | 1 | 0 |
| 7 | Blind assessors | 1 | 0 | 0 | 0 | 0 | 0 | 1 | 0 | 0 | 0 | 1 |
| 8 | Adequate follow-up | 1 | 1 | 1 | 1 | 1 | 1 | 0 | 1 | 1 | 1 | 1 |
| 9 | Intention-to-treat analysis | 0 | 0 | 1 | 1 | 1 | 1 | 0 | 1 | 0 | 0 | 0 |
| 10 | Between-group comparisons | 1 | 0 | 0 | 0 | 1 | 0 | 1 | 0 | 1 | 1 | 1 |
| 11 | Point estimates and variability | 1 | 1 | 1 | 1 | 1 | 0 | 1 | 1 | 1 | 1 | 1 |
|  | **Total** | **6** | **2** | **4** | **4** | **5** | **3** | **5** | **4** | **3** | **7** | **7** |
| 466 |  |  |  |  |  |  |  |  |  |  |  |  |
| 467 | *Not included in the final score |  |  |  |  |  |  |  |  |  |  |  |

## Discussion

To the best of our knowledge, this is the first systematic examination of BCI-driven robotic systems specific for hand rehabilitation. Through undertaking this review we found several limitations present from the studies identified and we examine these in more detail here and provide recommendations for future work in this area.

To provide clarity on the state of the current development of BCI-hand robot systems, we looked into the maturity of technology used in each study as determined by its readiness level (TRL). All but one in the prototype group was rated as having TRL 3 while the clinical group was more varied in their TRL (ranging from 5-7). The system used by Witkowski et al. [101], a prototype study, was rated TRL 4 due to the study being performed on the basis of improving and assessing its safety features. It is also worth noting that while a formal safety assessment was not performed for the TRL 3 prototypes of Stan et al. [94], Randazzo et al. [90] and Tacchino et al. [91], safety considerations and/or implementations were made; a criterion to be satisfied before proceeding to TRL 4. The system used by Chowdhury et al. is a good example of improving a TRL from 5 to 6 with a pilot clinical study published within the same year [78,79]. The two systems used in the RCT studies by Ang et al. [75] and Frolov et al. [80] achieved the highest score (TRL 7) among all of the studies which also meant that no BCI-hand robot system for stroke rehabilitation has ever been registered and commercially-released to date. This suggests that such systems lack the strong evidence that would propel commercialisation and technology adoption.

Heterogeneity in the study designs was apparent in both the clinical and prototype groups. The lack of control groups and random allocation in clinical studies (e.g., only 2 out of 7 studies are in the huge sample size RCT stage) made us unable to perform a meta-analysis of effects and continue the study by Cervera et al [51] with a focus on BCI-hand robot interventions. Results from the methodological quality assessment showed that only two studies [83,84] had a score of 7 in the PEDro scale. Although non-conclusive, these results support the notion that most of the studies are not aligned with the criteria of high-quality evidence-based interventions.

Almost all the clinical studies (except for Carino-Escobar et al. [85] and Frolov et al. [80]) limited their recruitment to chronic stroke patients. The reason may be due to the highly variable rates of recovery in patients at different stages in their disease [112]. Baseline treatments were also not reported among the clinical studies. Instead, the BCI-robot interventions were compared to control groups using standard arm therapy; an example of this was done by Ang et al. [75]. The heterogeneity of experimental designs reported in this review raises the need to develop clearly defined protocols when conducting BCI-hand robot studies on stroke patients. Until new systems have been assessed on this standard, it will be difficult to generate strong evidence supporting the effectiveness of BCI-robotic devices for hand rehabilitation.

In the development of any BCI-robotic device there are several design and feature considerations that need to be made to ensure that the systems are both fit for purpose and acceptable to the end-user. These design considerations must go beyond the scope of understanding the anatomy of the hand and the physiology of motor recovery in response to therapy. Feedback from stroke patients should also be an essential part of this design process. Among the extracted studies, we surveyed the extent of end-user involvement in the initial stages of development (i.e., through consultations, interviews and therapy observations) and we found that there were no explicit statements about these in the reports. We recommend, as good practice, for future work in this area to report the type and degree of patient and/or physician involvement in device development to allow reviewers and readers to more readily gauge the potential usability of the system.

We were able to profile the BCI-hand robot systems regarding their technical specifications and design features. In hardware terms, a BCI-hand robot system involves three major components: (1) An EEG data acquisition system with several electrodes connected to a signal amplifier; (2) A computer where raw EEG data is received then processed by filters and classifiers and where most of the cues and feedback during training is presented via a visual display; (3) a robotic hand rehabilitation system for providing the physical therapy back to the user.

The majority of the studies (N=19) used a BCI solely based on EEG while the rest were combined with other sensors: EEG with EMG [75,78,87,91,95–98], EEG with force sensors [79] and an EEG-EMG-EOG hybrid system [89,101]. The purpose of this integration is mainly to improve signal quality by accounting for artifact or to provide added modalities. Action potentials such as those caused by ocular, muscular and facial movements interfere with nearby electrodes and the presence of an added electrophysiological sensor accounting for these would enable the technician to perform noise cancellation techniques as a first step in signal processing.

The choice of EEG system as well as the type of electrodes provides a technical trade-off and affects the session both in terms of subjective experiences (i.e., ease-of-use, preparation time, cleaning, comfortability) and data performance. Due to the presence of a conducting gel/solution, standard “wet” electrodes provide a degree of confidence in preventing signal disruption within a short duration usually enough for a standard stroke therapy session. However, this also makes the setup, use and cleaning in the experiment more challenging, non-ambulatory and reliant on a specialised laboratory setup [10]. Conversely, dry electrodes offer an accessible, user-friendly and portable alternative by using dry metal pins or coatings that comb through hair and come in contact directly with the scalp. The signal fidelity of dry electrodes is still a matter of debate in the BCI community. A systematic comparison between dry passively-amplified and wet actively-amplified electrodes reported similar performance in the detection of event-related potentials (ERP) [113]. However, for a study involving dry active electrodes [114], high inter-electrode impedance resulted in increased single-trial and average noise levels as compared to both active and passive wet electrodes. In classifying MI, movement-related artifacts adversely affect active dry electrodes, but these can be addressed through a hybrid system of other physiological sensors to separate sources [115].

Almost all of the studies included used a standard EEG system with “wet” electrodes (e.g., g.USBamp by g.tec and BrainAmp by Brain Products) while three used Emotiv EPOC+, a semi-dry EEG system that uses sponge conductors infused with saline solution. While the use of dry electrodes has been observed in pilot and prototype studies of BCI-hand robot systems [67,64,93,102] and other motor imagery experiments [116–119], no dry EEG system was used in the final 30 studies that tested healthy or stroke participants. It is expected that as dry EEG systems continue to improve, their use in clinical studies of BCI will also become increasingly prominent.

The degree of BCI-robotic control for the majority of the studies (N=26) was limited to triggering the device to perform grasping (opening and closing of hand) and pinching (a thumb-index finger pinch or a 3-point thumb-index-middle finger pinch) movements using MI and other techniques. A triggered assistance strategy provides the minimum amount of active participation from the patient in a BCI-robot setup [37]. The main advantages of this is that it is easy to implement; requiring less computational complexity in signal processing. However, a higher spatial or temporal volitional control over the therapeutic device increases its functionality and can be used to develop more engaging tasks for the stroke therapy. Among the studies, no robotic control setup was able to perform digit-specific MI which corresponds to the spatial aspects of volitional control. This is a limitation caused by the non-invasive setup of EEG and is due to the low spatial resolution brought by the distances between electrodes [120]. The homunculus model, a representation of the human body in the motor strip, maps the areas of the brain where activations have been reported to occur for motor processes. The challenge of decoding each finger digit MI in one hand is that they only tend to occupy a small area in this strip. Hence even the highest resolution electrode placement system (i.e., the five percent or 10-5 system – up to 345 electrodes) would have difficulties accounting for digit-specific MI for BCI. In contrast to EEG, electrocorticography (ECoG) have been used to detect digit-specific MI. The electrodes of ECoG come in contact directly with the motor cortex and is an invasive procedure; making it non-ideal for use in BCI therapy [121].

It is worth noting however that some studies were successful in implementing continuous control based on ERD/ERS patterns. A continuous control strategy increases the temporal volitional control over the robot as opposed to triggered assistance where a threshold is applied, and the robot finishes the movement for the participant. Bundy et al. [77] and Norman et al. [82] were both able to apply continuous control of a 3-DOF pinch-grip exoskeleton based on spectral power while Bauer et al. [97] provided ERD-dependent control of finger extension for an end-effector robot. These continuous control strategies have been shown to be very useful in BCI-hand robots for assistive applications (i.e., partial or full device dependence for performing ADL tasks [122]). Whether this type of control can significantly improve stroke recovery is still in question as the strategy of robots for stroke rehabilitation can be more classified as a therapeutic “exercise” device.

Signal processing and machine learning play a vital role in the development of any EEG-based BCI. The pre-processing techniques (e.g., filtering, artifact removal), types of features computed from EEG, and the classifier used in machine learning can significantly affect the performance of the robotic system in classifying the user’s intent via MI [123]. False classification, especially during feedback, could be detrimental to the therapy regime as it relates to the reward and punishment mechanisms that are important in motor relearning [124]. For example, false negatives hinder the reward strategy that is essential to motivate the patient while false positives would also reward the action with the wrong intent. In this review, a critical appraisal of the signal processing techniques was done on each system to recognise the best practices involved. The current list of studies has revealed that approaches to develop MI-based EEG signal processing are highly diverse in nature, which makes it difficult to compare across the systems and hinders the development of new BCI systems informed by the strengths and weaknesses of existing state-of-the-art systems. The diversity in the design process can be beneficial to develop complex MI EEG-based BCI systems to achieve high efficiency and efficacy. However, such newly developed systems should be open sourced and easily reproducible by the research community to provide valid performance comparisons and drive forward the domain of robotic-assisted rehabilitation.

In addition to MI, other strategies for robotic control were reported. Diab et al. [103] and King et al. [104] both facilitated the movements of their respective orthoses by physical practice while Stan et al. [94] utilised a P-300 evoked potential speller BCI, where the user visually focused on a single alphanumerical character situated in a grid. The chosen character then corresponded to a command for the hand orthosis thereby producing the desired stimulus for the patient. While the latter study reported 100% accuracy rate in terms of intention and execution, the EEG channels were situated in the visual cortex rather than the motor strip which deviates from the goal of activating the desired brain region for plasticity. This highlights a broader issue on the intent behind a BCI-robotic system. Given that any potential signal that can be reliably modulated by a patient can be used to trigger a robot, and that such an approach would be antithetical to the goal of many MI-based systems, engineers may consider how they can tailor their systems to ensure that the appropriate control strategy (and corresponding neural networks) are implemented by a user (e.g. by taking a hybrid approach that includes EMG and force sensors).

In order to facilitate hand MI and account for significant time-points in the EEG data, all the studies employed a cue-feedback strategy during their trials. 19 of the studies presented a form of visual cue while the rest, except for two unspecified [84,102], involved cues in auditory (“bleep”) [91,95–98], textual [93,94,104] or verbal [103] forms. As for the provision of a matching sensory feedback, 16 studies presented a combination of kinaesthetic and visual feedback with some also providing auditory feedback during successful movement attempts. All the studies provided kinaesthetic feedback through their robotic devices. Some systems with visual feedback, such as Wang et al. [83], Li et al. [88], Chowdhury et al. in both of their clinical studies [78,79] and Ono et al. in their clinical [81] and pilot testing experiments [100], used a video of an actual hand performing the desired action. Ang et al. [75] and Stan et al. [94], in a different strategy, provided visual feedback through photo manipulation and textual display, respectively. While these two studies reported promising results, it should also be considered that such cue and feedback types (including Graz visualisations and auditory forms) are non-representative of hand movement and may not provide the same stimuli as an anthropomorphic representation of a hand moving its desired course. This may be essential when we base principles of stroke recovery in alignment with how MI correlates with AO – an underlying theme of the motor simulation theory proposed by Jeannerod [36]. Figure 4 shows how different kinds of visual cue and feedback can be presented to participants to help facilitate MI.

**Figure 4.**
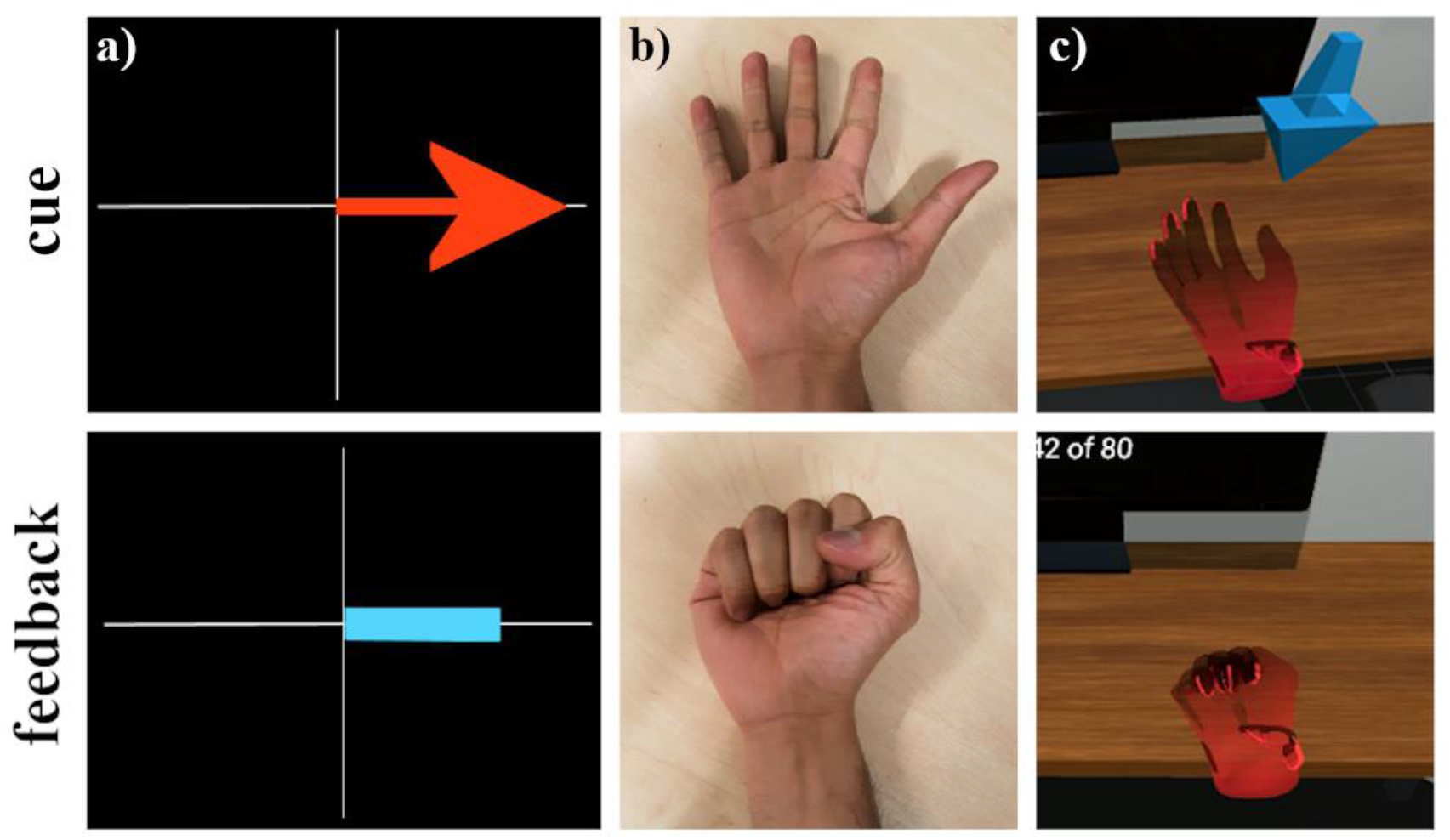
Visual cue and feedback during MI trials in different conditions. (a) Graz MI visualisations, (b) video recordings of hand movement and (c) virtual hand representation through VR/AR

### Future Directions

There is clearly great potential for the use of BCI-hand robots in the rehabilitation of an affected hand following stroke. Nevertheless, it is important to emphasise that there is currently insufficient evidence to support the use of such systems within clinical settings. Moreover, the purported benefits of these systems rest on conjectures that require empirical evidence. In other words, there are grounds for supposing that MI could be useful within these rehabilitation settings but no supporting evidence. This systematic review has also revealed that there are a number of technological limitations to existing BCI-hand robotic systems. We stress an urgent need to address these limitations to ensure that the systems meet the minimum required levels of product specification (in measuring brain activity, processing signals, delivering forces to the hand and providing rich feedback and motivating settings). We question the ethics or usefulness of conducting clinical trials with such systems until they can demonstrate minimum levels of technological capability. We consider below what standards these systems should obtain before subjecting them to a clinical trial and discuss might constitute an acceptable standard for a clinical trial.

### Ideal Setup for a BCI-hand Robot

We summarise the information revealed via the systematic review about what constitutes an acceptable setup for a BCI-hand robot for stroke rehabilitation. We focus on improving individual components in data acquisition, data processing, the hand rehabilitation robot, and the visual cue and feedback environment. Table 6 presents the features and specifications of a fully integrated acceptable system.

**Table 6.** Exemplary Features and Specifications of Future BCI-Hand Robot Systems.

| Component | Features and Specifications |
| --- | --- |
| Data Acquisition System and Software | • Dry EEG system with 8-16 channels, comfortable and easy to use<br>• Inclusion of other bio-signal sensors such as EMG, EOG, force, accelerometers to remove artifacts and improve classification<br>• Robust and reliable signal processing software: machine learning-based algorithms that discriminate brain states such as MI or evoked potentials with high classification accuracies ($\geq 95\%$) and lower calibration times |
| Hand Robot | • Safe, comfortable and aligned with the hand's range of motion<br>• Effective in providing kinaesthetic feedback<br>• Use of back-drivable or soft actuators that effectively assist movement without additional injury<br>• Multiple levels of safety and emergency features (mechanical, electronic, software), clear and obvious operation |
| Visual Cue and Feedback | • Provide rich visual cue and feedback to intended tasks, geometric representation of the hand (video or simulated environment), can be in multiple platforms such as display monitors or VR/AR on a head-mounted device<br>• Gamification of therapy exercises to provide an engaging regime to stroke patients |

The implementation of these features in an ideal BCI-robot setup needs to be weighed against socioeconomic factors in healthcare delivery for it to be considered market ready. An ideal BCI system should primarily provide above chance-level classification after the first session on the first day of therapy. Ideally, the classification algorithm should also translate and adapt to following sessions or days; reducing the number of training sessions and focusing on the main therapy tasks. An alternative approach is to focus on making the setup an engaging experience. In other words, the delivery of intervention can be started immediately when the patient wears the EEG cap and runs the BCI system. For the hand robot system, more straightforward criteria can be followed with the existence of the numerous design protocols, regulation standards and assessment matrices mentioned in this review. Nevertheless, end-user involvement in the design with the prioritisation of safety while allowing the most natural hand movement and ROM as possible is the recommended goal.

### Ideal Setup for Clinical Trials

We also propose a set of specialised criteria for BCI-hand robot systems in addition to the standard motor improvement scores (e.g. ARAT, FMA-UE) evaluated during clinical trials. Firstly, classification accuracies between intended and interpreted actions from the data acquisition and software component should always be accounted to track the effectiveness of BCI in executing the clinical task. In addition to this, system calibration and training procedures, especially its duration, should be detailed in the protocol to document the reliability of the classification algorithm. There is not much to consider in the use of robotic devices as they are most likely to be mature (if not yet commercially available) before being used as the hardware component in the study. However, the devices’ functionality (i.e., task to be performed, degree of control and motion, actuation and power transmission etc.) should always be stated as they contribute to the evaluation of interactions between other components in the system. Lastly, controls for the clinical study must always be included, even with small-scale patient studies. As discussed in this article, these controls may be in the form of sham, standard arm therapy (SAT), standard robotic therapy, congruency feedback and quality of stimuli among others. Having regarded and implemented these criteria would help homogenise the clinical data for future meta-analyses, strengthen evidence-based results and provide a reliable way of documentation for individual and/or interacting components.

### Proposed roadmap

We suggest that the immediate focus for BCI-controlled robotic device research should be around the engineering challenges. It is only when these challenges have been met that it is useful and ethical to subject the systems to clinical trials. We recommend that the challenges be broken down into the following elements: (1) data acquisition; (2) signal processing and classification; (3) robotic device; (4) priming and feedback environment; (5) integration of these four elements. The nature of these challenges means that a multidisciplinary approach is required (e.g. the inclusion of psychologists, cognitive neuroscientists and physiologists to drive the adoption of reliable neural data acquisition). It seems probable that progress will be made by different laboratories tackling some or all of these elements and coordinating information sharing and technology improvements. Once the challenges have been met (i.e. there is a system that is able to take neural signals and use these to help drive a robotic system capable of providing appropriate forces to the hand within a motivating environment) then robust clinical trials can be conducted to ensure that the promise of this approach does translate into solid empirical evidence supporting the use of these systems within clinical settings.

## Data Availability

A full database of selected studies including those extracted during the search and selection process is available from the corresponding author on reasonable request.

## Declarations

### Authors’ Contributions

PDEB and ECS performed the initial search and screening of the studies. MA performed the analysis of signal processing techniques. AA, AEJ and RJH made contribution to the robotics, design and other engineering aspects of the current work. FM provided analysis related to EEG and BCI. MMW contributed to the clinical and overall direction of the review. PDEB, FM and MMW were the major contributors to the writing of the manuscript. All authors read and approved the final manuscript.

### Ethics Approval and Consent to Participate

Not applicable.

### Consent for Publication

Not applicable.

## Acknowledgements

Not applicable.

## List of Abbreviations

ADL: Activities of Daily Living
ANN: Artificial Neural Network
AO: Action Observation
AR: Augmented Reality
ARAT: Action Research Arm Test
BCI: Brain-Computer Interface
BMI: Brain-Machine Interface
CDRH: Center for Devices and Radiological Health
CNN: Convolutional Neural Network
CSD: Covariate Shift Detection
CSP: Common Spatial Pattern
DC: Direct Current
ECoG: Electrocorticography
EEG: Electroencephalography
EMG: Electromyography
EOG: Electrooculography
ERD: Event-Related Desynchronisation
ERP: Event-Related Potential
ERS: Event-Related Synchronisation
ERSP: Event-Related Spectral Perturbation
FBCSP: Filter Bank Common Spatial Pattern
FIR: Finite Impulse Response
FMA-UE: Fugl-Meyer Motor Assessment – Upper Extremity
GS: Grip Strength
IIR: Infinite Impulse Response
LDA: Linear Discriminant Analysis
ME: Motor Execution
MI: Motor Imagery
PEDro: Physiotherapy Evidence Database
PMA: Premarket Approval
PRISMA: Preferred Reporting Items for Systematic Reviews and Meta-Analysis
PROSPERO: International Prospective Register of Systematic Reviews
PS: Pinch Strength
QSR: Quality System Requirement
RCT: Randomised Clinical Trial
RNN: Recurrent Neural Network
SAT: Standard Arm Therapy
SVM: Support Vector Machine
TRA: Technology Readiness Assessment
TRL: Technology Readiness Levels
UK-MRC: United Kingdom Medical Research Council
VR: Virtual Reality

## References

1. McHugh G, Swain ID. A comparison between reported therapy staffing levels and the department of health therapy staffing guidelines for stroke rehabilitation: a national survey. BMC Health Services Research. 2014;14:216.

2. Ntsiea MV. Current stroke rehabilitation services and physiotherapy research in South Africa. S Afr J Physiother. 2019;75:475–475.

3. Manguerra MV, Baniqued PDE, Abad AC, Baldovino RG, Dungao JR, Bugtai NT. Active motor control for an upper extremity exoskeleton. Advanced Science Letters. 2018;24:9937–8840.

4. Wolpaw JR, Wolpaw EW. Brain-Computer Interfaces, principles and practise. Oxford University Press USA; 2012.

5. Vidal JJ. Toward direct brain-computer communication. Annual review of biophysics and bioengineering. 1973;2:157–80.

6. Berger H. Über das elektrenkephalogramm des menschen. Archiv für Psychiatrie und Nervenkrankheiten. 1929;527–70.

7. Hochberg LR, Serruya MD, Friehs GM, Mukand JA, Saleh M, Caplan AH, et al. Neuronal ensemble control of prosthetic devices by a human with tetraplegia. Nature. 2006;442:164.

8. Silvoni S, Ramos-Murguialday A, Cavinato M, Volpato C, Cisotto G, Turolla A, et al. Brain-computer interface in stroke: a review of progress. Clin EEG Neurosci. 2011;42:245–52.

9. Birbaumer N, Ghanayim N, Hinterberger T, Iversen I, Kotchoubey B, Kubler A, et al. A spelling device for the paralysed. Nature. 1999;398:297–8.

10. Graimann B, Allison B, Pfurtscheller G. Brain–computer interfaces: a gentle introduction. In: Graimann B, Pfurtscheller G, Allison B, editors. Brain-Computer Interfaces: Revolutionizing Human-Computer Interaction [Internet]. Berlin, Heidelberg: Springer Berlin Heidelberg; 2010. p. 1–27. Available from: https://doi.org/10.1007/978-3-642-02091-9_1

11. Molteni F, Gasperini G, Cannaviello G, Guanziroli E. Exoskeleton and end-effector robots for upper and lower limbs rehabilitation: narrative review. PM&R. 2018;10:S174–88.

12. European Commission. Medical devices: guidance document - classification of medical devices [Internet]. United Kingdom Medicines and Healthcare products Regulation Agency; 2016 [cited 2018 Dec 10]. Available from: http://ec.europa.eu/growth/sectors/medical-devices/guidance/

13. Guiochet J, Hoang QAD, Kaaniche M, Powell DJ. Applying existing standards to a medical rehabilitation robot: limits and challenges. IROS. 2012.

14. Yue Z, Zhang X, Wang J. Hand rehabilitation robotics on poststroke motor recovery. Hindawi Behavioural Neurology. 2017;

15. Houwink A, Nijland RH, Geurts AC, Kwakkel G. Functional recovery of the paretic upper limb after stroke: Who regains hand capacity? Archives of physical medicine and rehabilitation. 2013;94:839–44.

16. Takahashi K, Domen K, Sakamoto T, Toshima M, Otaka Y, Seto M, et al. Efficacy of upper extremity robotic therapy in subacute poststroke hemiplegia: an exploratory randomized trial. Stroke. 2016;47:1385–8.

17. Fregni F, Pascual-Leone A. Hand motor recovery after stroke: tuning the orchestra to improve hand motor function. Cogn Behav Neurol. 2006;19:21–33.

18. Hallett M. Plasticity of the human motor cortex and recovery from stroke. Brain Res Brain Res Rev. 2001;36:169–74.

19. Zeiler SR, Krakauer JW. The interaction between training and plasticity in the poststroke brain. Curr Opin Neurol. 2013;26:609–16.

20. Mawase F, Uehara S, Bastian AJ, Celnik P. Motor learning enhances use-dependent plasticity. J Neurosci. 2017;37:2673–85.

21. Teplan M. Fundamentals of EEG measurement. Measurement Science Review. 2002;2.

22. Cohen MX. Analyzing neural time series data: theory and practice [Internet]. Cambridge; 2014. Available from: http://lib.ugent.be/catalog/rug01:002161004

23. Li M, Xu G, Xie J, Chen C. A review: motor rehabilitation after stroke with control based on human intent. Proc Inst Mech Eng H. 2018;232:344–60.

24. Pfurtscheller G, Lopes da Silva FH. Event-related EEG/MEG synchronization and desynchronization: basic principles. Clin Neurophysiol. 1999;110:1842–57.

25. Jiang X, Bian GB, Tian Z. Removal of artifacts from EEG signals: a review. Sensors. 2019;19.

26. Mammone N, Morabito FC. Enhanced automatic wavelet independent component analysis for electroencephalographic artifact removal. Entropy. 2014;16:6553–72.

27. Val-Calvo M, Alvarez-Sanchez JR, Ferrandez-Vicente JM, Fernandez E. Optimization of real-time EEG artifact removal and emotion estimation for human-robot interaction applications. Frontiers in Computational Neuroscience. 2019;

28. Kilicarslan A, Vidal JLC. Characterization and real-time removal of motion artifacts from EEG signals. Journal of Neural Engineering. 2019;16.

29. Chang CY, Hsu SH, Pion-Tonachini L, Jung TP. Evaluation of artifact subspace reconstruction for automatic artifact components removal in multi-channel EEG recordings. IEEE Transactions on Biomedical Engineering. 2019;

30. Lotte F, Guan C. Regularizing common spatial patterns to improve BCI designs: unified theory and new algorithms. IEEE Transactions on Biomedical Engineering. 2011;58:355–62.

31. Roy Y, Banville H, Albuquerque I, Gramfort A, Falk TH, Faubert J. Deep learning-based electroencephalography analysis: a systematic review. Journal of Neural Engineering. 2019;16.

32. Schirrmeister RB, Springenberg JT, Fiederer LDJ, Glasstetter M, Eggensperger K, Tangermann M, et al. Deep learning with convolutional neural networks for EEG decoding and visualization. Human Brain Mapping. 2017;38:5391–420.

33. Hardwick RM, Caspers S, Eickhoff SB, Swinnen SP. Neural correlates of action: comparing meta-analyses of imagery, observation, and execution. Neuroscience and Biobehavioral Reviews. 2018;31–44.

34. Xu L, Zhang H, Hui M, Long Z, Jin Z, Liu Y, et al. Motor execution and motor imagery: a comparison of functional connectivity patterns based on graph theory. Neuroscience. 2014;261:184–94.

35. Sharma N, Baron JC. Does motor imagery share neural networks with executed movement: a multivariate fMRI analysis. Front Hum Neurosci. 2013;7:564–564.

36. Jeannerod M. Neural simulation of action: a unifying mechanism for motor cognition. NeuroImage. 2001;14:S103–9.

37. Marchal-Crespo L, Reinkensmeyer DJ. Review of control strategies for robotic movement training after neurologic injury. Journal of NeuroEngineering and Rehabilitation. 2009;6.

38. Brookes J, Mushtaq F, Jamieson E, Fath AJ, Bingham G, Culmer P, et al. Exploring disturbance as a force for good in motor learning. PLOS ONE. 2020;15.

39. Mulder T. Motor imagery and action observation: cognitive tools for rehabilitation. J Neural Transm (Vienna). 2007;114:1265–78.

40. Vogt S, Di Rienzo F, Collet C, Collins A, Guillot A. Multiple roles of motor imagery during action observation. Frontiers in Human Neuroscience. 2013;7:807.

41. Friesen CL, Bardouille T, Neyedli HF, Boe SG. Combined action observation and motor imagery neurofeedback for modulation of brain activity. Frontiers in Human Neuroscience. 2017;10:692.

42. Eaves DL, Riach M, Holmes PS, Wright DJ. Motor imagery during action observation: a brief review of evidence, theory and future research opportunities. Frontiers in Neuroscience. 2016;10:514.

43. Kim T, Frank C, Schack T. A systematic investigation of the effect of action observation training and motor imagery training on the development of mental representation structure and skill performance. Frontiers in Human Neuroscience. 2017;11:499.

44. Nakano H, Kodama T. Motor imagery and action observation as effective tools for physical therapy. Neurological Physical Therapy. IntechOpen; 2017.

45. Laver KE, Lange B, George S, Deutsch JE, Saposnik G, Crotty M. Virtual reality for stroke rehabilitation. Cochrane Database Syst Rev. 2017;11:CD008349–CD008349.

46. Brookes J, Warburton M, Alghadier M, Mon-Williams M, Mushtaq F. Studying human behavior with virtual reality: the Unity experiment framework. Behavior Research Methods [Internet]. 2019; Available from: https://doi.org/10.3758/s13428-019-01242-0

47. Vourvopoulos A, Cardona JEM, Badia SB i. Optimizing motor imagery neurofeedback through the use of multimodal immersive virtual reality and motor priming. 2015 International Conference on Virtual Rehabilitation (ICVR). 2015. p. 228–34.

48. Johnston M, Bonetti D, Joice S, Pollard B, Morrison V, Francis JJ, et al. Recovery from disability after stroke as a target for a behavioural intervention: results of a randomized controlled trial. Disability and Rehabilitation. 2007;29:1117–27.

49. Hülsmann F, Frank C, Senna I, Ernst MO, Schack T, Botsch M. Superimposed skilled performance in a virtual mirror improves motor performance and cognitive representation of a full body motor action. Frontiers in Robotics and AI. 2019;6:43.

50. Monge-Pereira E, Ibanez-Pereda J, Alguacil-Diego IM, Serrano JI, Spottorno-Rubio MP, Molina-Rueda F. Use of electroencephalography brain-computer interface systems as a rehabilitative approach for upper limb function after a stroke: a systematic review. PM R. 2017;9:918–32.

51. Cervera MA, Soekadar SR, Ushiba J, Millan J d. R, Liu M, Birbaumer N, et al. Brain-computer interfaces for post-stroke motor rehabilitation: a meta-analysis. bioRxiv [Internet]. 2017; Available from: http://biorxiv.org/content/early/2017/11/24/224618.abstract

52. McConnell AC, Moioli RC, Brasil FL, Vallejo M, Corne DW, Vargas PA, et al. Robotic devices and brain-machine interfaces for hand rehabilitation post-stroke. Journal of Rehabilitation Medicine. 2017;49:449–60.

53. Fugl-Meyer AR, Jaasko L, Leyman I, Olsson S, Steglind S. The post-stroke hemiplegic patient. 1. a method for evaluation of physical performance. Scand J Rehabil Med. 1975;7:13–31.

54. Lyle RC. A performance test for assessment of upper limb function in physical rehabilitation treatment and research. International Journal of Rehabilitation Research [Internet]. 1981;4. Available from: https://journals.lww.com/intjrehabilres/Fulltext/1981/12000/A_performance_test_for_assessment_of_upper_limb.1.aspx

55. Matthews WB. Aids to the examination of the peripheral nervous system. Journal of the Neurological Sciences. 1977;33:299.

56. Office of the Director of Defense Research and Engineering Washington DC. Technology Readiness Assessment (TRA) deskbook [Internet]. Fort Belvoir, VA, USA: Defense Technical Information Center; 2009 Jul. Report No.: ADA524200. Available from: https://apps.dtic.mil/dtic/tr/fulltext/u2/a524200.pdf

57. Maher CG, Sherrington C, Herbert RD, Moseley AM, Elkins M. Reliability of the PEDro scale for rating quality of randomized controlled trials. Physical Therapy. 2003;83:713–21.

58. PEDro Scale [Internet]. 1999 [cited 2018 Nov 30]. Available from: www.pedro.org.au/english/downloads/pedro-scale/

59. Belardinelli P, Laer L, Ortiz E, Braun C, Gharabaghi A. Plasticity of premotor cortico-muscular coherence in severely impaired stroke patients with hand paralysis. Neuroimage Clin. 2017;14:726– 33.

60. Bergamasco M, Frisoli A, Fontana M, Loconsole C, Leonardis D, Troncossi M, et al. Preliminary results of BRAVO project: brain computer interfaces for robotic enhanced action in visuo-motor tasks. IEEE Int Conf Rehabil Robot. 2011;2011:5975377.

61. Broetz D, Braun C, Weber C, Soekadar SR, Caria A, Birbaumer N. Combination of brain-computer interface training and goal-directed physical therapy in chronic stroke: a case report. Neurorehabil Neural Repair. 2010;24:674–9.

62. Cincotti F, Pichiorri F, Arico P, Aloise F, Leotta F, de Vico Fallani F, et al. EEG-based brain-computer interface to support post-stroke motor rehabilitation of the upper limb. Conf Proc IEEE Eng Med Biol Soc. 2012;2012:4112–5.

63. George K, Iniguez A, Donze H, Kizhakkumthala S. Design, implementation and evaluation of a brain-computer interface controlled mechanical arm for rehabilitation. 2014 IEEE International Instrumentation and Measurement Technology Conference (I2MTC) Proceedings. 2014. p. 1326–8.

64. Ushiba J, Morishita A, Maeda T. A task-oriented brain-computer interface rehabilitation system for patients with stroke hemiplegia. 2014 4th International Conference on Wireless Communications, Vehicular Technology, Information Theory and Aerospace & Electronic Systems (VITAE). 2014. p. 1–3.

65. Rathee D, Chowdhury A, Meena YK, Dutta A, McDonough S, Prasad G. Brain–machine interface-driven post-stroke upper-limb functional recovery correlates with beta-band mediated cortical networks. IEEE Transactions on Neural Systems and Rehabilitation Engineering. 2019;27:1020–31.

66. Naros G, Gharabaghi A. Physiological and behavioral effects of beta-tACS on brain self-regulation in chronic stroke. Brain Stimul. 2017;10:251–9.

67. Norman S. Brain computer interface design for robot assisted neurorehabilitation [Internet]. University of California, Irvine; 2017. Available from: https://escholarship.org/uc/item/4v18v0d3

68. Pellegrino G, Tomasevic L, Tombini M, Assenza G, Bravi M, Sterzi S, et al. Inter-hemispheric coupling changes associate with motor improvements after robotic stroke rehabilitation. Restor Neurol Neurosci. 2012;30:497–510.

69. Bousseta R, El Ouakouak I, Gharbi M, Regragui F. EEG based brain computer interface for controlling a robot arm movement through thought. IRBM. 2018;39:129–35.

70. Formaggio E, Storti SF, Boscolo Galazzo I, Gandolfi M, Geroin C, Smania N, et al. Modulation of event-related desynchronization in robot-assisted hand performance: brain oscillatory changes in active, passive and imagined movements. J Neuroeng Rehabil. 2013;10:24.

71. Sarasola-Sanz A, Irastorza-Landa N, Lopez-Larraz E, Bibian C, Helmhold F, Broetz D, et al. A hybrid brain-machine interface based on EEG and EMG activity for the motor rehabilitation of stroke patients. IEEE Int Conf Rehabil Robot. 2017;2017:895–900.

72. Shiman F, Irastorza-Landa N, Sarasola-Sanz A, Spuler M, Birbaumer N, Ramos-Murguialday A. Towards decoding of functional movements from the same limb using EEG. Conf Proc IEEE Eng Med Biol Soc. 2015;2015:1922–5.

73. Muralidharan A, Chae J, Taylor DM. Extracting attempted hand movements from EEGs in people with complete hand paralysis following stroke. Front Neurosci. 2011;5:39.

74. Ono T, Mukaino M, Ushiba J. Functional recovery in upper limb function in stroke survivors by using brain-computer interface: a single case A-B-A-B design. Conf Proc IEEE Eng Med Biol Soc. 2013;2013:265–8.

75. Ang KK, Guan C, Phua KS, Wang C, Zhou L, Tang KY, et al. Brain-computer interface-based robotic end effector system for wrist and hand rehabilitation: results of a three-armed randomized controlled trial for chronic stroke. Frontiers in Neuroengineering. 2014;7:30–30.

76. Barsotti M, Leonardis D, Loconsole C, Solazzi M, Sotgiu E, Procopio C, et al. A full upper limb robotic exoskeleton for reaching and grasping rehabilitation triggered by MI-BCI. 2015 IEEE International Conference on Rehabilitation Robotics (ICORR). 2015. p. 49–54.

77. Bundy DT, Souders L, Baranyai K, Leonard L, Schalk G, Coker R, et al. Contralesional brain-computer interface control of a powered exoskeleton for motor recovery in chronic stroke survivors. Stroke. 2017;48:1908–15.

78. Chowdhury A, Raza H, Meena YK, Dutta A, Prasad G. Online covariate shift detection based adaptive brain-computer interface to trigger hand exoskeleton feedback for neuro-rehabilitation. IEEE Transactions on Cognitive and Developmental Systems. 2018;1–1.

79. Chowdhury A, Meena YK, Raza H, Bhushan B, Uttam AK, Pandey N, et al. Active physical practice followed by mental practice using BCI-driven hand exoskeleton: a pilot trial for clinical effectiveness and usability. IEEE J Biomed Health Inform. 2018;22:1786–95.

80. Frolov AA, Mokienko O, Lyukmanov R, Biryukova E, Kotov S, Turbina L, et al. Post-stroke rehabilitation training with a motor-imagery-based brain-computer interface (BCI)-controlled hand exoskeleton: a randomized controlled multicenter trial. Frontiers in Neuroscience. 2017;11:400–400.

81. Ono Y, Tominaga T, Murata T. Digital mirror box: an interactive hand-motor BMI rehabilitation tool for stroke patients. 2016 Asia-Pacific Signal and Information Processing Association Annual Summit and Conference (APSIPA). 2016. p. s1–7.

82. Norman SL, McFarland DJ, Miner A, Cramer SC, Wolbrecht ET, Wolpaw JR, et al. Controlling pre-movement sensorimotor rhythm can improve finger extension after stroke. Journal of Neural Engineering. 2018;15:056026.

83. Wang X, Wong W, Sun R, Chu WC, Tong KY. Differentiated effects of robot hand training with and without neural guidance on neuroplasticity patterns in chronic stroke. Frontiers in Neurology. 2018;9:810.

84. Tsuchimoto S, Shindo K, Hotta F, Hanakawa T, Liu M, Ushiba J. Sensorimotor connectivity after motor exercise with neurofeedback in post-stroke patients with hemiplegia. Neuroscience. 2019;416:109–25.

85. Carino-Escobar RI, Carillo-Mora P, Valdes-Cristerna R, Rodriguez-Barragan MA, Hernandez-Arenas C, Quinzaños-Fresnedo J, et al. Longtitudinal analysis of stroke patient’ brain rhythms during an interview with a brain-computer interface. Neural Plasticity. 2019;2019:11.

86. Cantillo-Negrete J, Carino-Escobar RI, Elias-Vinas D, Gutierrez-Martinez J. Control signal for a mechatronic hand orthosis aimed for neurorehabilitation. 2015 Pan American Health Care Exchanges (PAHCE). 2015. p. 1–4.

87. Chowdhury A, Raza H, Dutta A, Nishad SS, Saxena A, Prasad G. A study on cortico-muscular coupling in finger motions for exoskeleton assisted neuro-rehabilitation. Conf Proc IEEE Eng Med Biol Soc. 2015;2015:4610–4.

88. Li M, He B, Liang Z, Zhao CG, Chen J, Zhuo Y, et al. An attention-controlled hand exoskeleton for the rehabilitation of finger extension and flexion using a rigid-soft combined mechanism. Frontiers in Neurorobotics. 2019;13:34.

89. Zhang J, Wang B, Zhang C, Xiao Y, Wang MY. An EEG/EMG/EOG-based multimodal human-machine interface to real-time control of a soft robot hand. Frontiers in Neurorobotics. 2019;13:7.

90. Randazzo L, Iturrate I, Perdikis S, Millán Jd.R. mano: a wearable hand exoskeleton for activities of daily living and neurorehabilitation. IEEE Robotics and Automation Letters. 2018;3:500–7.

91. Tacchino G, Gandolla M, Coelli S, Barbieri R, Pedrocchi A, Bianchi AM. EEG analysis during active and assisted repetitive movements: evidence for differences in neural engagement. IEEE Trans Neural Syst Rehabil Eng. 2017;25:761–71.

92. Coffey AL, Leamy DJ, Ward TE. A novel BCI-controlled pneumatic glove system for home-based neurorehabilitation. Conf Proc IEEE Eng Med Biol Soc. 2014;2014:3622–5.

93. Holmes CD, Wronkiewicz M, Somers T, Liu J, Russell E, Kim D, et al. IpsiHand Bravo: an improved EEG-based brain-computer interface for hand motor control rehabilitation. Conf Proc IEEE Eng Med Biol Soc. 2012;2012:1749–52.

94. Stan A, Irimia DC, Botezatu NA, Lupu RG. Controlling a hand orthosis by means of P300-based brain computer interface. 2015 E-Health and Bioengineering Conference (EHB). 2015. p. 1–4.

95. Ramos-Murguialday A, Schürholz M, Caggiano V, Wildgruber M, Caria A, Hammer EM, et al. Proprioceptive feedback and brain computer interface (BCI) based neuroprostheses. PLOS ONE. 2012;7:e47048.

96. Ramos-Murguialday A, Birbaumer N. Brain oscillatory signatures of motor tasks. J Neurophysiol. 2015;113:3663–82.

97. Bauer R, Fels M, Vukelic M, Ziemann U, Gharabaghi A. Bridging the gap between motor imagery and motor execution with a brain-robot interface. Neuroimage. 2015;108:319–27.

98. Naros G, Naros I, Grimm F, Ziemann U, Gharabaghi A. Reinforcement learning of self-regulated sensorimotor beta-oscillations improves motor performance. Neuroimage. 2016;134:142–52.

99. Vukelic M, Gharabaghi A. Oscillatory entrainment of the motor cortical network during motor imagery is modulated by the feedback modality. Neuroimage. 2015;111:1–11.

100. Ono Y, Wada K, Kurata M, Seki N. Enhancement of motor-imagery ability via combined action observation and motor-imagery training with proprioceptive neurofeedback. Neuropsychologia. 2018;114:134–42.

101. Witkowski M, Cortese M, Cempini M, Mellinger J, Vitiello N, Soekadar S. Enhancing brain-machine interface (BMI) control of a hand exoskeleton using electrooculography (EOG). Journal of NeuroEngineering and Rehabilitation. 2014;11:165.

102. Fok S, Schwartz R, Wronkiewicz M, Holmes C, Zhang J, Somers T, et al. An EEG-based brain computer interface for rehabilitation and restoration of hand control following stroke using ipsilateral cortical physiology. Conf Proc IEEE Eng Med Biol Soc. 2011;2011:6277–80.

103. Diab MS, Hussain Z, Mahmoud S. Restoring function in paralyzed limbs using EEG. 2016 IEEE 59th International Midwest Symposium on Circuits and Systems (MWSCAS). 2016. p. 1–4.

104. King CE, Wang PT, Mizuta M, Reinkensmeyer DJ, Do AH, Moromugi S, et al. Noninvasive brain-computer interface driven hand orthosis. Conf Proc IEEE Eng Med Biol Soc. 2011;2011:5786– 9.

105. Ang KK, Chin ZY, Wang C, Guan C, Zhang H. Filter bank common spatial pattern algorithm on BCI competition IV datasets 2a and 2b. Frontiers in neuroscience. 2012;6:39–39.

106. Bobrov PD, Korshakov AV, Roshchin VI, Frolov AA. Bayesian classifier for brain-computer interface based on mental representation of movements. Zh Vyssh Nerv Deiat Im I P Pavlova. 2012;62:89–99.

107. Gonzalez CDV, Azuela JHS, Espino ER, Ponce VH. Classification of motor imagery imagery EEG signals with CSP filtering through neural networks models. Advances in Soft Computing, Lecture Notes in Computer Science. Springer, Cham; 2018. p. 123–35.

108. Chaudhari R, Galiyawala HJ. A review on motor imagery signal classification for BCI. Signal Processing: An International Journal. 2017;11:16–34.

109. Oña ED, Cano-de la Cuerda R, Sanchez-Herrera P, Balaguer C, Jardon A. A review of robotics in neurorehabilitation: towards an automated process for upper limb. J Healthc Eng. 2018;

110. Kersten P. Principles of physiotherapy assessment and outcome measures. Physical Management in Neurological Rehabilitation. 2004. p. 29–46.

111. Dovat L, Lambercy O, Ruffieux Y, Chapuis D, Gassert R, Bleuler H, et al. A haptic knob for rehabilitation after stroke. 2006 IEEE/RSJ International Conference on Intelligent Robots and Systems. Beijing, China: IEEE; 2006.

112. Ballester BR, Maier M, Duff A, Cameirao M, Bermudez S, Duarte E, et al. A critical time window for recovery extends beyond one-year post-stroke. Journal of Neurophysiology. 2019;350–7.

113. Kam JWY, Griffin S, Shen A, Patel S, Hinrichs H, Heinze HJ, et al. Systematic comparison between a wireless EEG system with dry electrodes and a wired EEG system with wet electrodes. NeuroImage. 2019;184:119–29.

114. Mathewson KE, Harrison TJL, Kizuk SAD. High and dry? Comparing active dry EEG electrodes to active and passive wet electrodes. Psychophysiology. 2017;54:74–82.

115. Saab J, Battes B, Grosse-Wentrup M. Simultaneous EEG recordings with dry and wet electrodes in motor-imagery. In proceedings. Graz, Austria; 2011.

116. Abdalsalam E, Yusoff MZ, Kamel N, Malik AS, Mahmoud D. Classification of four class motor imagery for brain computer interface. In: Ibrahim H, Iqbal S, Teoh SS, Mustaffa MT, editors. 9th International Conference on Robotic, Vision, Signal Processing and Power Applications. Springer Singapore; 2017. p. 297–305.

117. Grummett TS, Leibbrandt RE, Lewis TW, DeLos Angeles D, Powers DMW, Willoughby JO, et al. Measurement of neural signals from inexpensive, wireless and dry EEG systems. Physiological Measurement. 2015;36.

118. Mladenov T, Kim K, Nooshabadi S. Accurate motor imagery based dry electrode brain-computer interface system for consumer applications. 2012 IEEE 16th International Symposium on Consumer Electronics. 2012. p. 1–4.

119. Guger C, Krausz G, Edliner G. Brain-computer interface control with dry EEG electrodes. Proceedings of the 5th International Brain-Computer Interface Conference. 2011. p. 316–9.

120. Srinivasan R. Methods to improve the spatial resolution of EEG. International Journal of Bioelectromagnetism. 1999;1:102–11.

121. Liao K, Xiao R, Gonzalez J, Ding L. Decoding individual finger movements from one hand using human EEG signals. PLOS ONE. 2014;9:e85192.

122. Johnson MJ, Micera S, Shibata T, Guglielmelli E. Rehabilitation and assistive robotics [TC Spotlight]. IEEE Robotics & Automation Magazine. 2008;15:16–110.

123. Arvaneh M, Guan C, Ang K, Quek C. Optimizing the channel selection and classification accuracy in EEG-based BCI. Biomedical Engineering, IEEE Transactions on. 2011;58:1865–73.

124. Quattrocchi G, Greenwood R, Rothwell JC, Galea JM, Bestmann S. Reward and punishment enhance motor adaptation in stroke. Journal of Neurology, Neurosurgery, and Psychiatry. 2016;88:730–6.

